# Circulating metabolites associated with psoriasis in the UK Biobank and the HUNT Study: A cross-sectional study of 270,848 participants

**DOI:** 10.1101/2025.10.28.25338943

**Authors:** Alya G. A. Arham, Abhibhav Sharma, Laurent F. Thomas, Ravi Ramessur, Julia Debik, Marion Denos, Mari Hoff, Vibeke Videm, Lavinia Paternoster, Catherine Smith, Bjørn O. Åsvold, Guro F. Giskeødegård, Ben M. Brumpton, Mari Løset

## Abstract

**Background:** Psoriasis is recognized as a systemic inflammatory disease associated with metabolic dysregulation. Understanding these metabolic changes may reveal biomarkers to elucidate disease mechanisms and predict comorbidities. While previous studies have identified psoriasis-associated metabolites, findings are often limited by sample sizes and lack validation.

**Objectives:** We aimed to identify circulating metabolites associated with psoriasis, including cutaneous activity, severity, and psoriatic arthritis. Further, we investigated whether the metabolic signature was disease-specific compared to other immune-mediated inflammatory diseases (IMIDs).

**Methods:** We performed a cross-sectional analysis of 270,848 White/European individuals from the UK Biobank (n=253,924) and HUNT (n=16,924). Both cohorts used nuclear magnetic resonance spectroscopy to quantify metabolite levels, covering lipoprotein fractions and subfractions, fatty acids, and small-molecular metabolites. For each metabolite, we performed multivariable linear regression adjusting for age, sex, BMI, smoking status, and use of lipid-lowering medications.

**Results:** The metabolomic profile of psoriasis was largely consistent across the two cohorts. In the model adjusted for age and sex, 116 metabolic measures were associated with psoriasis in both cohorts. After full adjustment, only Glycoprotein acetyls (GlycA) remained associated with psoriasis (coefficient [95% CI]: 0.09 [0.07-0.11] in UK Biobank and 0.11 [0.06-0.17] in HUNT). Despite more substantial metabolic alterations in cutaneous-active psoriasis, GlycA was also elevated in HUNT participants reporting no active psoriasis rash (coefficient [95% CI]: 0.12 [0.04-0.20] in non-cutaneous-active and 0.11 [0.03-0.19] in cutaneous-active psoriasis). In HUNT, severe psoriasis exhibited more pronounced metabolic alterations compared to non-severe psoriasis. Across both cohorts, phenylalanine levels were highly elevated in psoriatic arthritis compared to cutaneous psoriasis (coefficient [95% CI]: 0.41 [0.20-0.62] in UK Biobank and 0.47 [0.28-0.67] in HUNT). All IMIDs showed elevated GlycA and reduced albumin, with milder changes in atopic dermatitis. Notably, psoriasis in the HUNT cohort exhibited a distinct lipoprotein profile compared to other IMIDs.

**Conclusions:** This large-scale, cross-cohort study confirms metabolic alterations in individuals with psoriasis and highlights elevated GlycA levels regardless of cutaneous activity. The distinct metabolomic profile of psoriasis relative to other IMIDs suggests a potentially unique systemic profile. These findings offer a foundation for advancing biomarker research and mechanistic studies for psoriasis.

**Graphical abstract:** 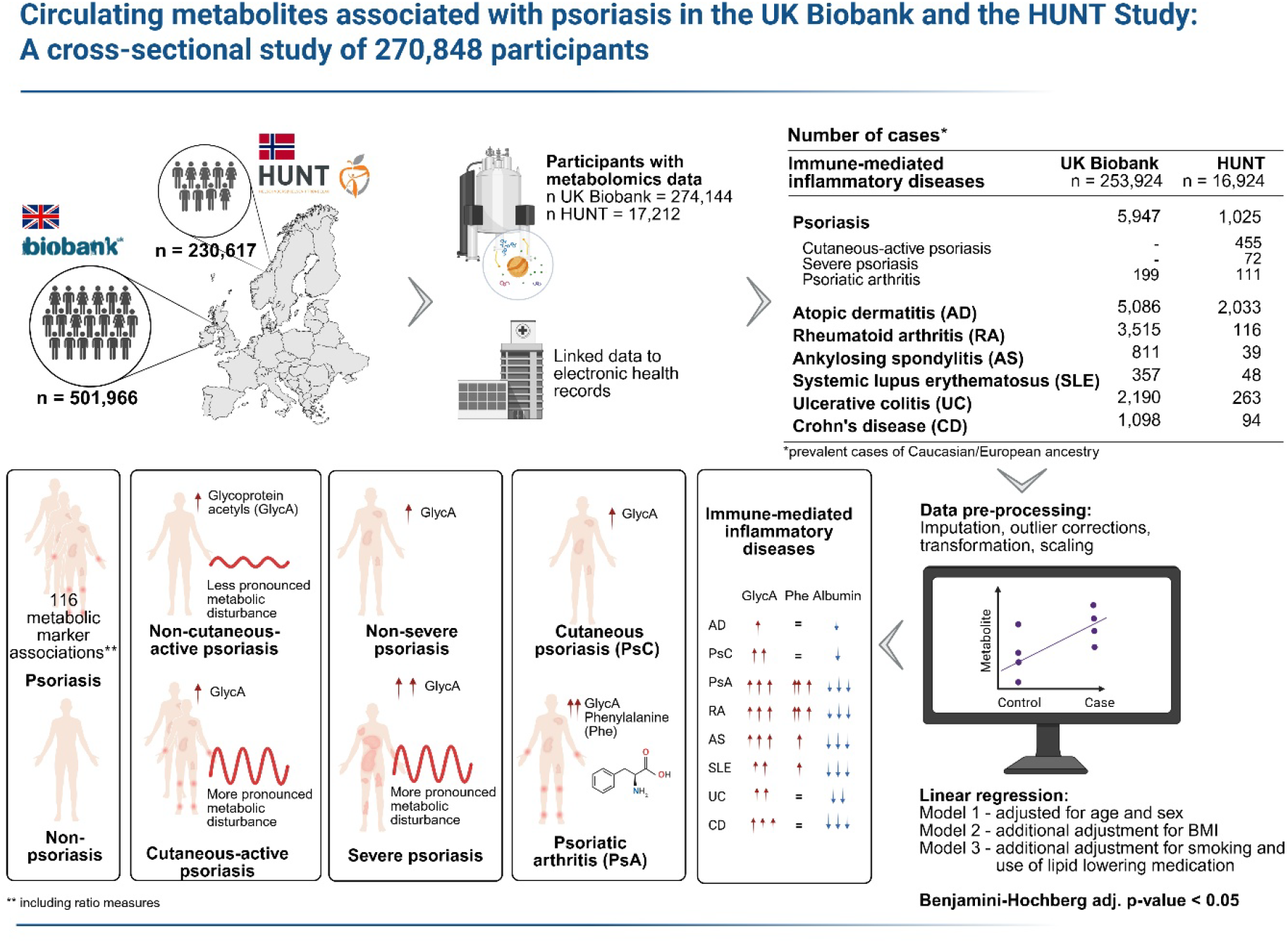

Created in BioRender, Arham A (2026) https://BioRender.com/o3aw61n licensed under CC BY 4.0.

## Introduction

Psoriasis is a common chronic immune-mediated inflammatory skin disease, caused by the interplay between multiple genetic predispositions and environmental triggers.^1^ People with psoriasis, particularly those with moderate to severe disease, are at increased risk of other medical conditions, including cardiometabolic diseases and depression.^2^ As there is no cure yet,^3^ current treatment strategies are primarily reactive, aiming to alleviate symptoms and prolong the duration of remission periods.^4^

In the past two decades, psoriasis has been increasingly recognized as a systemic disease with significant metabolic dysregulations.^5^ Although the full extent of its systemic metabolic effects remains poorly understood, several studies have investigated circulating metabolites associated with psoriasis, in both plasma and serum samples.^6^ Common metabolites have emerged across studies, including amino acids, carnitines, and fatty acids.^7^ However, these studies are often limited by small sample sizes and lack validation, raising concerns about the robustness of the results^8^ and hindering the translational impact.

In this study, we aimed to investigate the associations between circulating metabolic markers and psoriasis in two large population-based cohorts—UK Biobank and The Trøndelag Health Study (HUNT). We also investigated the associations of the metabolic markers with several key clinical features of psoriasis, including cutaneous disease activity, characterized by skin rashes and remission; disease severity; and psoriatic arthritis (PsA). Further, we explored shared metabolic signatures between psoriasis and other immune-mediated inflammatory diseases (IMIDs), including atopic dermatitis (AD), rheumatoid arthritis (RA), ankylosing spondylitis (AS), systemic lupus erythematosus (SLE), ulcerative colitis (UC), and Crohn’s disease (CD).

### Participants and methods

#### Study populations

##### UK Biobank

UK Biobank is a population-based prospective study conducted in the United Kingdom between 2006 and 2010.^9,10^ It comprises around 500,000 participants, aged between 40 and 69 years at recruitment. The study collected extensive data, covering sociodemographic, environmental and lifestyle factors, alongside health status, physical measurements, and biological samples, including blood, urine, and saliva. Participant data have also been linked to medical records, including hospital records, general practitioner data, and death registry. In this study, we included only individuals of White ethnicity to improve the comparability between the two cohorts.

##### HUNT

HUNT is an ongoing population-based cohort study in Trøndelag County, Norway, which has been conducted in four surveys over the past 40 years: HUNT1 (1984–1986), HUNT2 (1995–1997), HUNT3 (2006–2008), and HUNT4 (2017–2019)^.11,12^ The surveys collected data covering clinical measurements, biological samples, and questionnaires on socioeconomic status and health-related information.^13^ Participant data have been linked to medical records and national health registries through their unique national identification numbers. In this study, we used data from HUNT3, comprising approximately 50,800 adults aged ≥20 years. We used genetic ancestry data to determine participant ethnicity,^14^ and only individuals of European ancestry were included.

#### Disease definitions

We defined all cases as participants who had received their diagnosis prior to blood sampling. In UK Biobank, we defined psoriasis, AD, RA, AS, SLE, UC, and CD using the ‘first occurrences of medical conditions’ data fields, generated based on the self-reported medical conditions, primary care, hospital inpatient, and death records. For PsA, we used the L40.5, M07.0, M07.1, M07.2, and M07.3 codes from the ICD-10 summary diagnosis data field. In HUNT, we identified psoriasis and AD cases using self-reported data and diagnostic codes from multiple sources: ICPC-2 for primary care, and ICD-9/10 for hospital records, reimbursement claims, and prescription records. We defined SLE, UC, and CD cases using the ICD-9/10 codes only. Meanwhile for PsA, RA, and AS, we only included rheumatologist-validated diagnoses^.15,16^ In both cohorts, we defined cutaneous-limited psoriasis (PsC) as non-PsA psoriasis cases.

In HUNT, we defined cutaneous-active psoriasis as self-reported psoriasis rash within two weeks prior to blood sampling, based on the HUNT3 psoriasis questionnaire. We defined severe psoriasis cases in HUNT by using the level of clinical intervention received up to one year prior to blood sampling, as suggested in a previous literature^17^. In UK Biobank, we could not define cutaneous activity and disease severity due to insufficient information. Full details on the disease definitions are provided in the **Supplementary Method S1 and S2**.

#### Covariates

We selected the covariates—age, sex, BMI, smoking status, and use of lipid-lowering medications— based on domain knowledge and previous literature.^18-23^ The complete list of the UK Biobank data fields and HUNT variables used as covariates is provided in the **Supplementary Method S3**. To harmonize smoking status between cohorts, in HUNT, we grouped occasional and daily smokers as “*current smokers*”. For lipid-lowering medication use, we used the self-reported medication data for UK Biobank, and the ATC code C10 in the Norwegian Prescribed Drug Registry for HUNT. In our dataset, missing values were present for some of the covariates (<1% in UK Biobank and ≤2.5% in HUNT). Therefore, we created an imputed dataset for each cohort using IterativeImputer, a multivariable imputation method in Python, inspired by the Multiple Imputation by Chained Equations (MICE) in R (https://scikit-learn.org/stable/modules/generated/sklearn.impute.IterativeImputer.html).

#### Metabolomics measurements

We used metabolomics data from 274,144 non-fasting plasma samples in UK Biobank (UK Biobank metabolomics phase 2 data release, July 2023) and 17,212 non-fasting serum samples in HUNT. Samples from both cohorts were analysed using high-throughput nuclear magnetic resonance (NMR) spectroscopy, provided by Nightingale Health Ltd., Helsinki, Finland. The full details on the quantification method have been previously described.^24^ The measurement covers 168 absolute metabolite concentrations and 81 metabolite ratios, including various low-molecular-weight metabolites, such as ketone bodies, amino acids, and glycolysis-related metabolites, as well as lipids and lipoprotein subfractions^.24,25^

#### Metabolomics data pre-processing

For UK Biobank, we corrected the measurements for technical variations using ukbnmr package, which applies a multistep robust linear regression to adjust each metabolite value for technical factors.^26^ For HUNT, we used previously corrected metabolite measures for differences between batches and type of NMR spectrometers.^27^ Before imputation, we excluded observations with more than 10% missing metabolite values. For the UK Biobank dataset, we also removed samples flagged for low protein levels which indicates potential dilution. We then performed k-Nearest Neighbor (kNN) imputation for values missing at random and Quantile Regression Imputation of Left-Censored data (QRILC) for values missing not at random. Details on the rationale for determining the type of missingness for the imputation are provided in the **Supplementary Method S4**. We replaced outliers, defined as values outside the four interquartile ranges from the median (4×IQR), with the corresponding 4×IQR threshold values. To facilitate comparability across metabolites, we applied log1p-transformation and standardized values to z-scores.

#### Statistical analysis

We performed multivariable linear regression for each measured metabolite, with disease status as the independent variable (binary) and the metabolite concentration as the dependent variable (continuous). To minimize bias in the associations, we adjusted for age and sex in model 1, included additional adjustment for BMI in model 2, and further accounted for smoking status and the use of lipid-lowering medications in model 3. For UK Biobank, we also adjusted for the assessment centre in all models. To account for non-normal distributions of residuals, we used heteroscedasticity-robust standard error estimation. We performed multiple testing correction using the Benjamini–Hochberg procedure, with statistical significance defined as adjusted p-value < 0.05. We performed the statistical analysis in Python, with codes provided on https://github.com/AlyaGhina/circulating-metabolomics-profile-of-psoriasis.

## Results

### Case numbers of the eligible participants

In this study, we ascertained a total of 270,848 individuals of White ethnicity/European ancestry with available metabolomics data from UK Biobank (n = 253,924) and HUNT (n = 16,924). Among these individuals, 6,972 had psoriasis prior to blood sampling (n UK Biobank = 5,947; n HUNT = 1,025). A chart showing the numbers of participants included in the analysis and the timepoints for defining psoriasis cases are provided in **Figure 1**.

**Figure 1.**
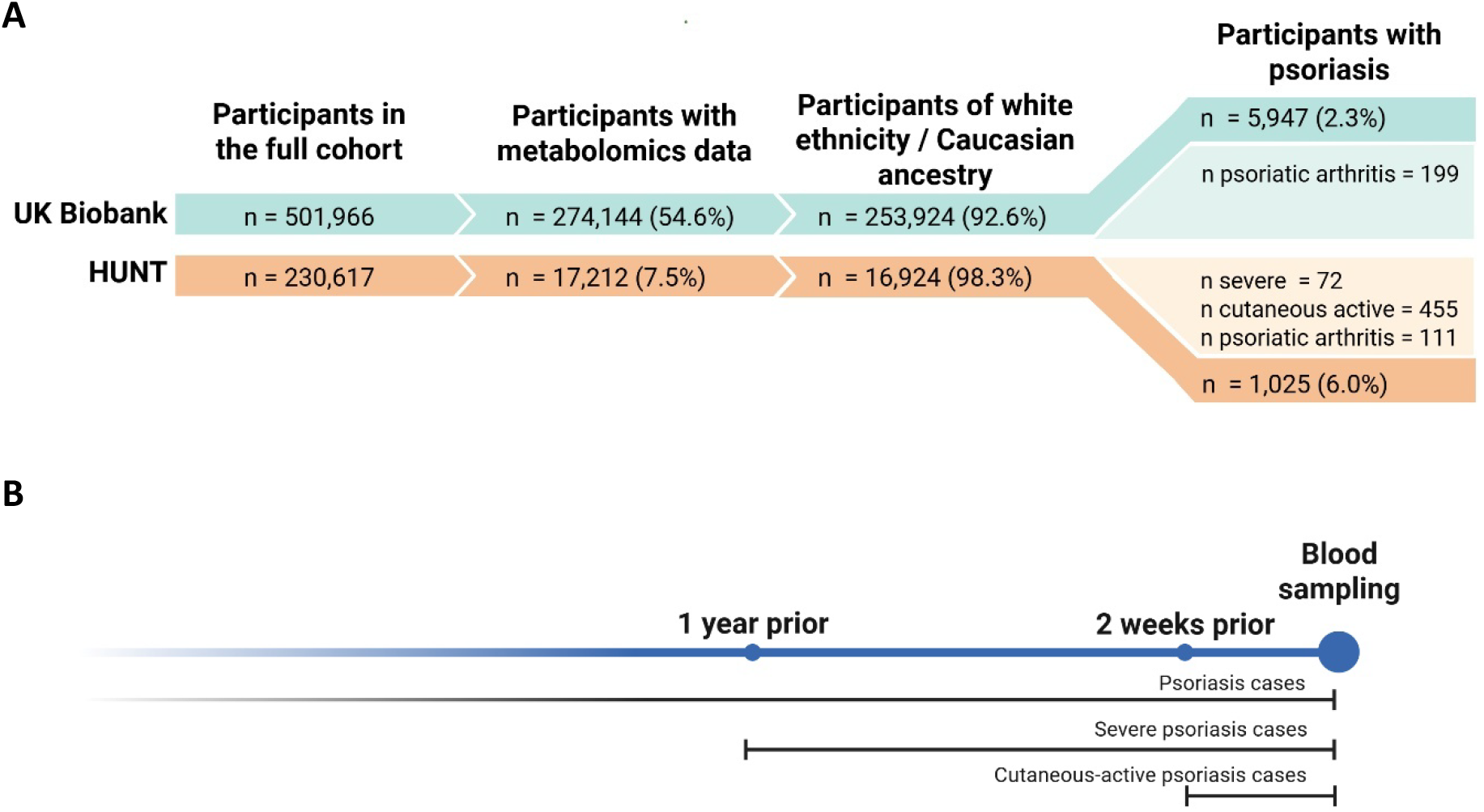
(A) Overview of the number of participants included in the analysis, along with the number of psoriasis cases, including severe psoriasis, cutaneous-active psoriasis, and psoriatic arthritis. The percentages indicate the proportions of participants relative to the previous filtering step. (B) Overview of the timepoints for defining psoriasis cases: psoriasis cases were participants who have had psoriasis diagnosis at any time point prior to blood sampling; severe cases were psoriasis cases who have had severe disease within one year prior to blood sampling; cutaneous-active cases were psoriasis cases who have had psoriasis rash within 2 weeks prior to blood sampling.

### General characteristics of the cohorts

Among White/European participants with available metabolomics data, the proportion of psoriasis cases was 2.3% (n = 5,947) in the UK Biobank and 6.0% in HUNT (n = 1,025). Overall, both cohorts had generally similar sex distributions (proportion of male participants: 46% in UK Biobank and in HUNT) and median BMI (26.8, IQR: [24.2, 29.9] kg/m^2^ in UK Biobank vs. 26.7 IQR: [24.2, 29.9] in HUNT). Compared to HUNT, the UK Biobank cohort had a higher median age (58.0 IQR: [50.0, 63.0] vs. 53.2 IQR: [40.8, 63.4] years) and fasting time (3.0 IQR: [2.0, 4.0] vs. 2.0 IQR: [1.0, 3.0] hours), a higher proportion of lipid-lowering medication users (17.7% vs. 12.1%), and fewer current smokers (10.4% vs. 24.0%). In both cohorts, individuals with psoriasis had a higher observed median BMI, and were more likely to smoke and use lipid-lowering medication compared to those without psoriasis. The characteristics of the participants with and without psoriasis in both cohorts are provided in **Table 1**.

**Table 1:** General characteristics of participants with and without psoriasis who had metabolomics data available in the UK Biobank and HUNT.

|  |  | UK Biobank<br>(n = 253,924) |  | HUNT<br>(n = 16,924) |  |
| --- | --- | --- | --- | --- | --- |
|  |  | No psoriasis<br>(n = 247,977) | Psoriasis<br>(n = 5,947) | No psoriasis<br>(n = 15,899) | Psoriasis<br>(n = 1,025) |
| Age (years),<br>median [Q1, Q3] |  | 58.0<br>[50.0, 63.0] | 58.0<br>[51.0, 63.0] | 53.0<br>[40.5, 63.4] | 56.1<br>[45.5, 63.4] |
| Sex, n (%) | Male | 113,695 (45.8) | 3,011<br>(50.6) | 7,341 (46.2) | 481 (46.9) |
|  | Female | 134,282 (54.2) | 2,936<br>(49.4) | 8,558 (53.8) | 544 (53.1) |
| BMI (kg/m<sup>2</sup>),<br>median [Q1, Q3] |  | 26.7<br>[24.2,29.9] | 27.5<br>[24.7,30.8] | 26.6<br>[24.1,29.5] | 27.5<br>[24.5,30.9] |
| Fasting time (hours),<br>median [Q1, Q3] |  | 3.0<br>[2.0,4.0] | 3.0<br>[3.0,4.0] | 2.0<br>[1.0,3.0] | 2.0<br>[1.0,3.0] |
| Smoking status, n (%) | Current | 25,638(10.3) | 804 (13.5) | 3,744 (23.5) | 316 (30.8) |
|  | Previous | 87,629 (35.3) | 2,497<br>(42.0) | 4,910 (30.9) | 365 (35.6) |
|  | Never | 134,710 (54.3) | 2,646<br>(44.5) | 6,834 (43.0) | 326 (31.8) |
| On lipid-lowering<br>medication, n (%) | No | 204,258 (82.4) | 4,674<br>(78.6) | 13997 (88.0) | 875 (85.4) |
|  | Yes | 43,719 (17.6) | 1,273<br>(21.4) | 1902 (12.0) | 150 (14.6) |

Abbreviations: Q1, Quartile 1; Q3: Quartile 3; BMI, Body Mass Index.

### Metabolite associations with psoriasis

In the model adjusted for age and sex only (model 1), 116 metabolic measures were associated with psoriasis in both cohorts, most showing consistent effect estimates (**Supplementary Figure S1** and **Supplementary Table S1**). These metabolic measures cover various metabolite groups, including those related to inflammation; amino acids including histidine and glutamine; glucose; fatty acids including saturated, monounsaturated, and total fatty acids; lipoprotein fractions and subfractions. However, these associations were attenuated after further adjustment for BMI (model 2), and even more so after additional adjustment for smoking status and the use of lipid-lowering medication (model 3) (**Supplementary Figures S2** and **S3**). In the fully adjusted model, only Glycoprotein acetyls (GlycA), an inflammatory marker, was associated with psoriasis in both cohorts (**Figure 2**).

**Figure 2.**
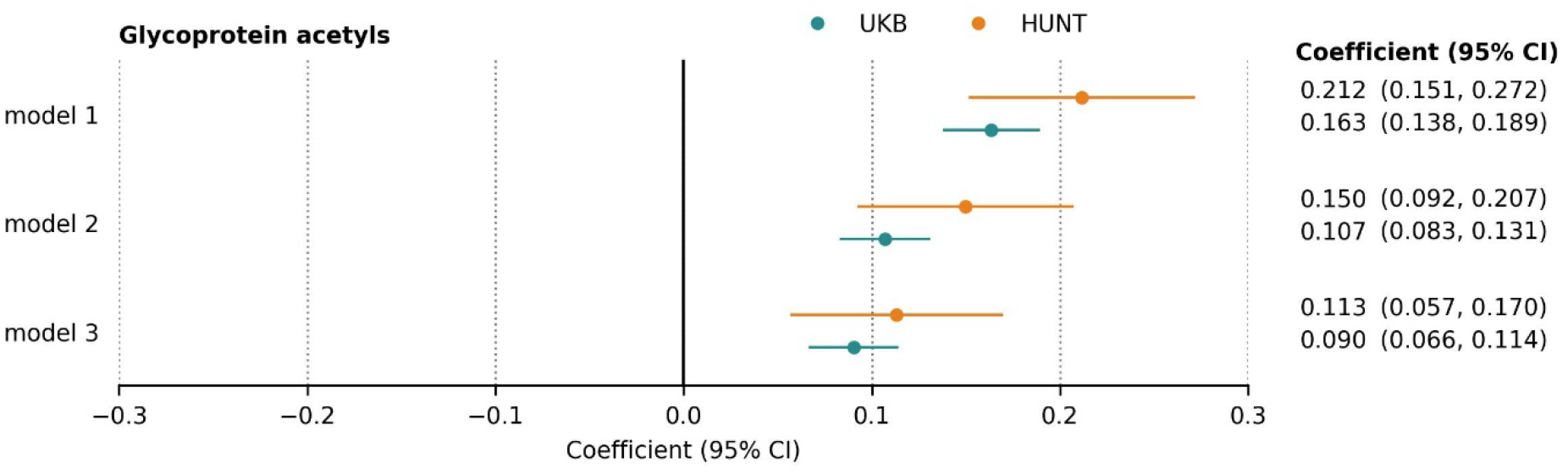
The association between Glycoprotein acetyls and psoriasis, in UK Biobank and HUNT, using models 1, 2, and 3. Model 1 was adjusted for age and sex; Model 2 for age, sex, and BMI; Model 3 for age, sex, BMI, smoking status, and use of lipid-lowering medication. Full results for all metabolites are provided in the **Supplementary Table S1**. CI = Confidence Interval.

We further examined metabolite associations with cutaneous activity, psoriasis severity and PsA. The characteristics of the participants with cutaneous-active, non-cutaneous-active, severe psoriasis, non-severe psoriasis, PsC, and PsA are provided in the **Supplementary Table S2**. Overall, we observed a trend of larger effect magnitudes in cutaneous-active psoriasis (**Figure 3A, Supplementary Figure S4, Supplementary Table S3**). Interestingly, GlycA levels were also elevated in individuals reporting non-cutaneous-active disease (coefficient [95% CI]: 0.12 [0.04-0.20] in non-cutaneous-active and 0.11 [0.03-0.19] in cutaneous-active psoriasis). Compared to non-severe psoriasis, severe psoriasis showed more pronounced metabolic alterations, particularly in the lipid profile, and showed a higher GlycA level elevation (**Figure 3B, Supplementary Figure S5, Supplementary Table S4**). When comparing PsC and PsA cases, we observed a significant elevation of phenylalanine levels in PsA cases in both cohorts (**Figure 4, Supplementary Table S5**). We then extended the analysis to investigate whether phenylalanine elevation is specific to PsA or reflects a broader association with inflammatory arthritis (**Figure 5**). In HUNT, phenylalanine was elevated across inflammatory arthritis cases, but not in PsC and other non-arthritis diseases. In UK Biobank, however, the elevation was consistent only for RA.

**Figure 3.**
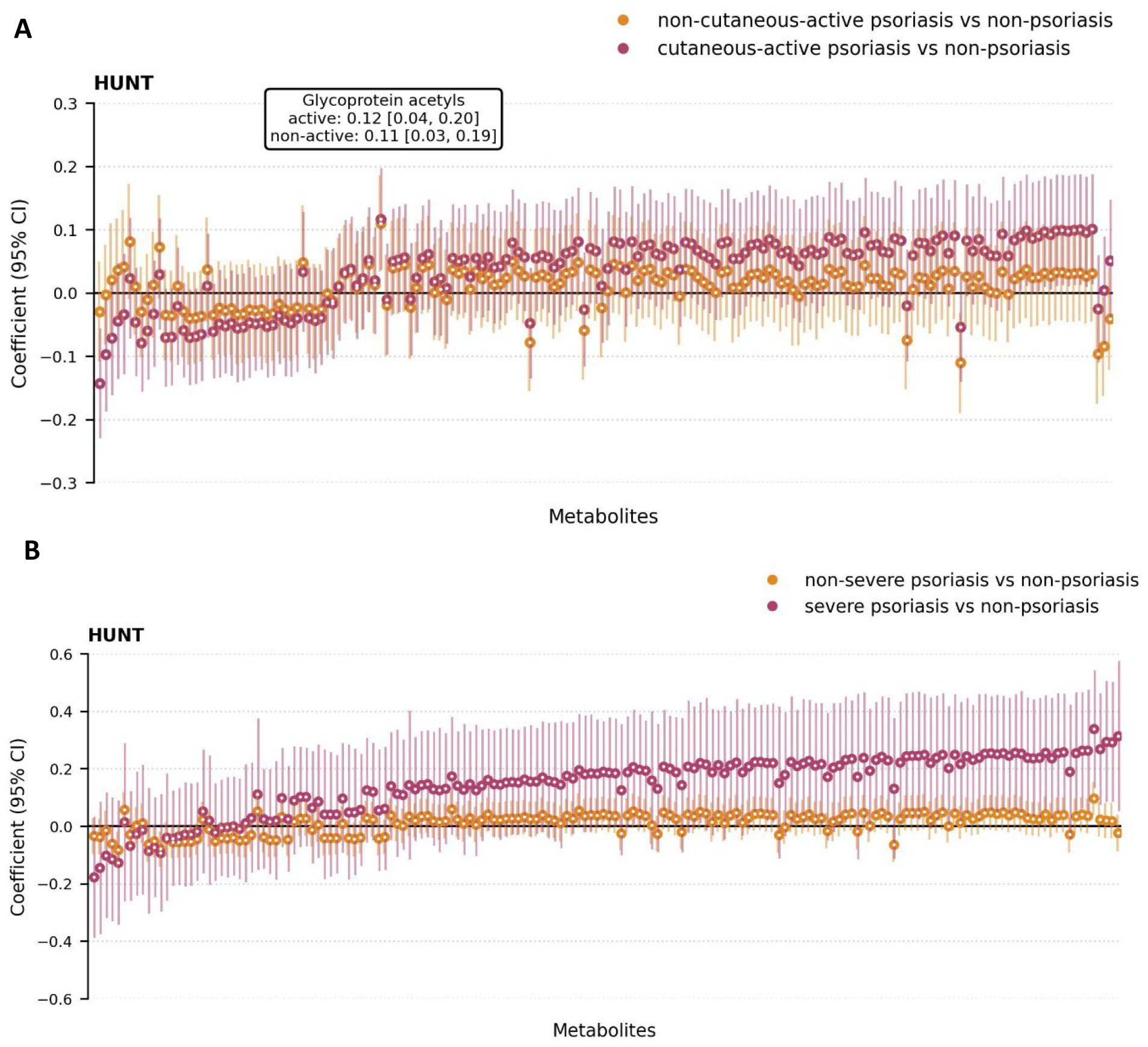
Forest plot showing the coefficients (95% CI) of the metabolites for non-cutaneous-active and cutaneous-active psoriasis (A) and for non-severe and severe psoriasis (B) in HUNT. Numbers of participants included in the analyses: n cutaneous-active psoriasis = 455, n non-cutaneous-active psoriasis = 570, n severe psoriasis = 72, n non-severe psoriasis= 953, n non-psoriasis = 15,899. Analyses were adjusted for age, sex, BMI, smoking status, and use of lipid-lowering medications (model 3). Adjusted p-values < 0.05 are indicated by solid fill. Full results are provided in the **Supplementary Table S1**. CI = Confidence Interval.

**Figure 4.**
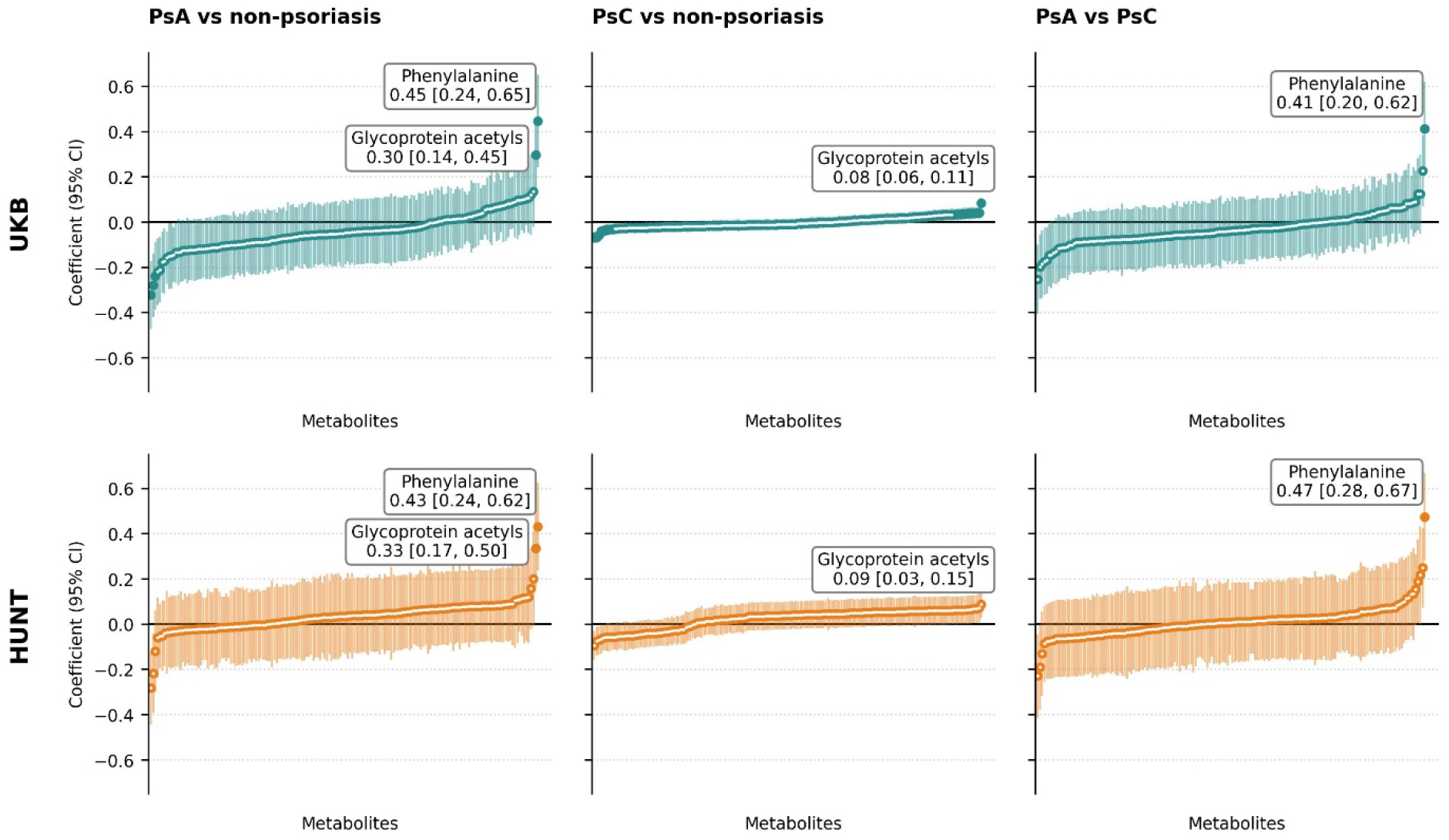
Forest plot showing the coefficients (95% CI) of the metabolites in PsA vs non-psoriasis (left), PsC vs non-psoriasis (middle), PsA vs PsC (right). Numbers of participants included in the analyses: n PsA in UKB = 199, n PsC in UKB = 5,748, n non-psoriasis in UKB = 247,977, n PsA in HUNT = 111, n PsC in HUNT = 914, n non-psoriasis in HUNT = 15,899. Analyses were adjusted for age, sex, BMI, smoking status, and use of lipid-lowering medications (model 3). Adjusted p-values < 0.05 are indicated by solid fill. Full results are provided in the **Supplementary Table S1.** UKB = UK Biobank, CI = Confidence Interval.

**Figure 5.**
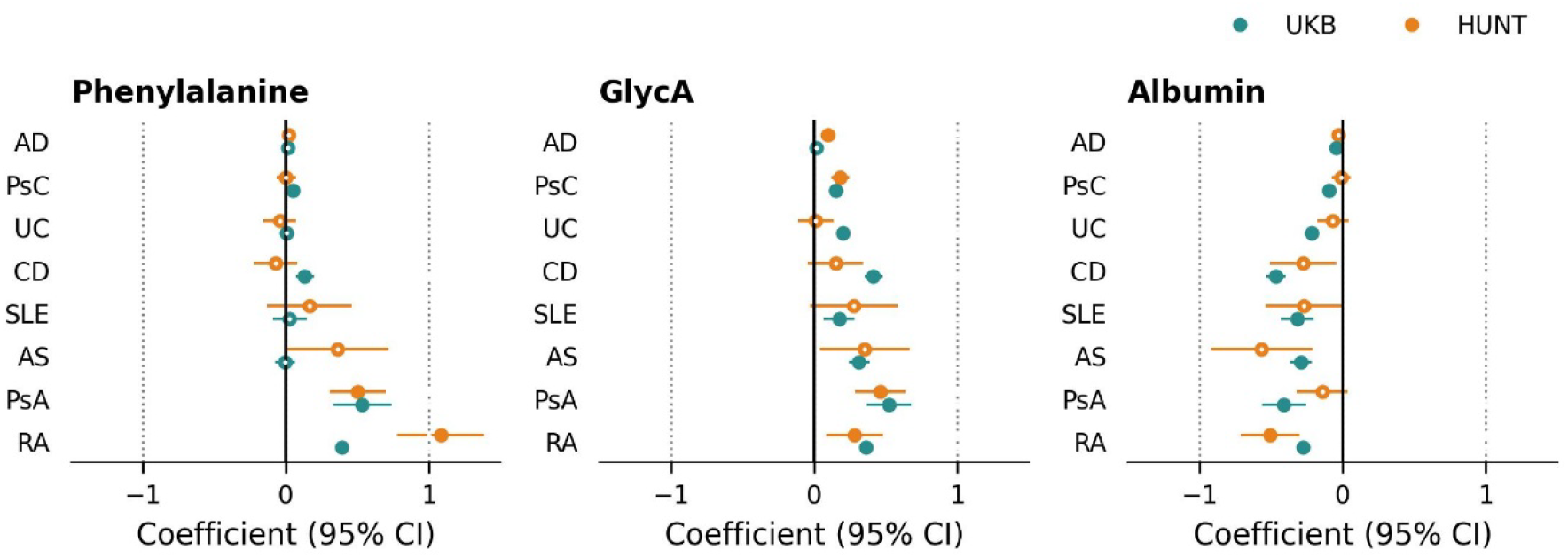
Forest plot showing the coefficients (95% CI) for phenylalanine, GlycA and albumin across all IMIDs in both cohorts. Analyses were performed using age- and sex-adjusted models. Adjusted p-values < 0.05 are indicated by solid fill. Full results are provided in the **Supplementary Table S1, S5, and S7.** UKB = UK Biobank, CI = Confidence Interval, AD = Atopic Dermatitis, PsC = Cutaneous-limited Psoriasis, PsA = Psoriatic Arthritis, AS = Ankylosing Spondylitis, RA = Rheumatoid Arthritis, SLE = Systemic Lupus Erythematosus, UC = Ulcerative Colitis, CD = Crohn’s Disease.

*Metabolite associations across IMIDs*To investigate shared and distinct metabolic features between psoriasis and other IMIDs, we compared the metabolic profile of psoriasis (PsC and PsA combined and separately) with profiles of AD, RA, AS, SLE, UC, and CD. The characteristics of the participants for each IMID in both cohorts are provided in the **Supplementary Table S6**. All IMIDs exhibited signs of inflammation as shown by elevated GlycA and depleted albumin levels (**Figure 5**). Based on the global metabolic profile across both cohorts, AD consistently showed milder metabolic alterations, followed by PsC, in contrast to the more pronounced alterations in PsA and other IMIDs. In HUNT, psoriasis, including both PsC and PsA, exhibited a distinct lipoprotein fraction and subfraction profile compared to the other IMIDs (**Figure 6, Supplementary Table S1, S5, and S7**), including IDL, LDL, VLDL, L-LDL, M-LDL, S-LDL, XS-VLDL, S-VLDL, M-VLDL, L-VLDL, XL-VLDL, and XXL-VLDL compositions. However, this pattern was not replicated in the UK Biobank.

**Figure 6.**
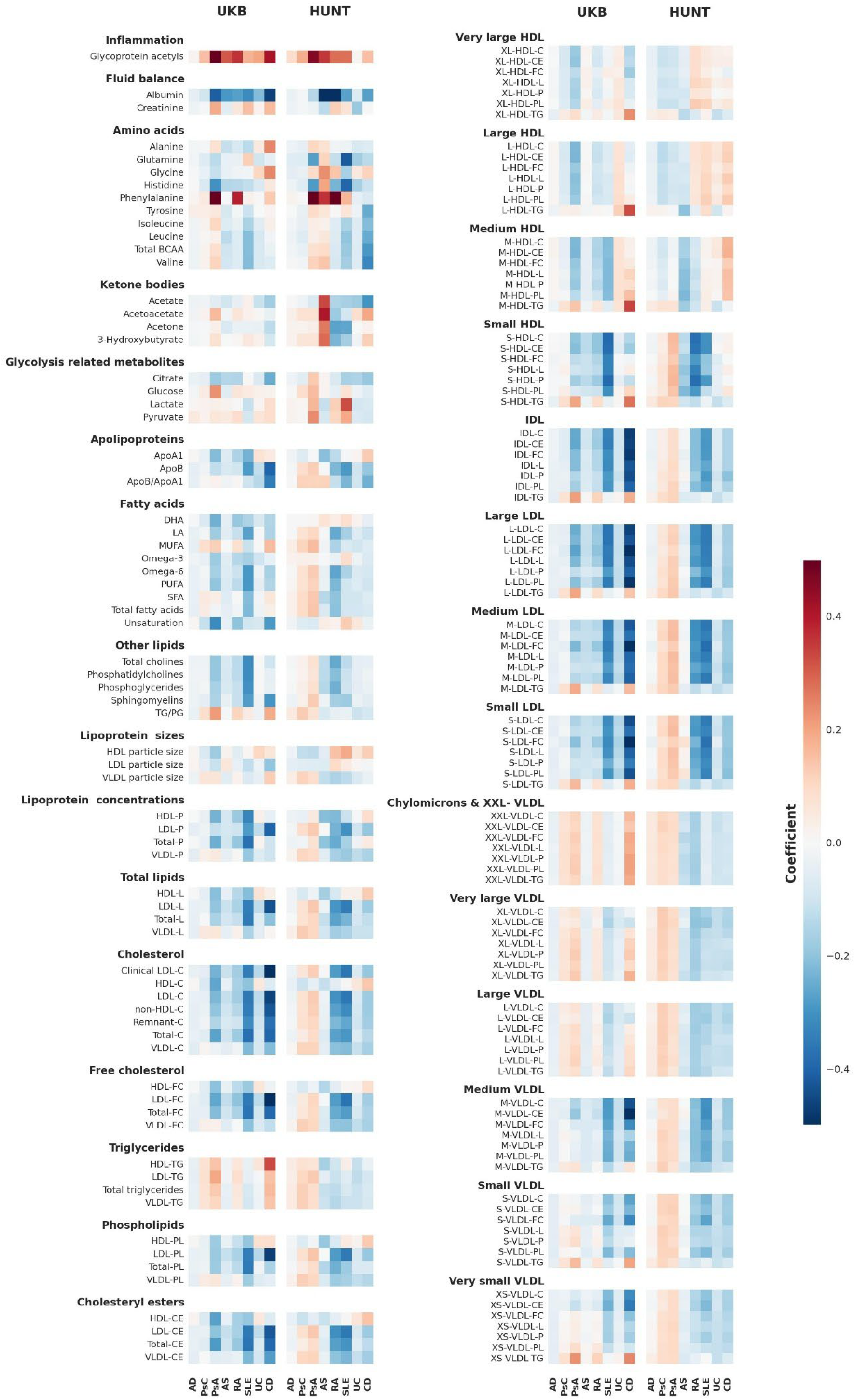
Heatmaps showing metabolite coefficients for different IMIDs in UK Biobank and HUNT, adjusted for age and sex. Each row represents a metabolite. The colour gradient indicates the magnitude of the coefficient, with darker colours representing larger magnitudes. The full description of the metabolite abbreviations is provided in **Supplementary Table S8.** AD = Atopic Dermatitis, PsC = Cutaneous-limited Psoriasis, PsA = Psoriatic Arthritis, AS = Ankylosing Spondylitis, RA = Rheumatoid Arthritis, SLE = Systemic Lupus Erythematosus, UC = Ulcerative Colitis, CD = Crohn’s Disease.

## Discussion

In this study, we investigated the associations between circulating NMR metabolites and psoriasis, PsA, and six other IMIDs, leveraging metabolomics data from two large population-based biobanks: UK Biobank and HUNT. In the age-and sex-adjusted model, 116 metabolic measures were found associated with psoriasis, and the overall metabolomic profiles were largely consistent across the two cohorts.

Notably, GlycA, a composite biomarker reflecting the glycosylation of multiple acute-phase proteins and low-grade chronic inflammation,^28-30^ emerged as the only metabolite associated with psoriasis in both cohorts after full adjustment for age, sex, BMI, smoking status, and use of lipid-lowering medications. While its association with psoriasis is expected, our findings add to the growing body of evidence supporting the systemic inflammatory nature of psoriasis^5,31,32^ Our observation also aligns with previous studies linking GlycA to psoriasis^33,34^ as well as other IMIDs, such as RA^35^ and SLE^36^.

Considering the varying cutaneous activity at the time of sampling, we assessed the associations separately for those who reported psoriasis rash within two weeks prior to blood sampling. Interestingly, we observed that GlycA level was also elevated in non-cutaneous-active psoriasis, suggesting that systemic inflammation persists even when visible skin symptoms subside. A recent review discusses the potential inflammatory memory mechanisms within psoriatic skin tissue,^37^ and our GlycA results support that the inflammatory effects may also occur beyond the skin.

Regarding disease severity, we observed that individuals with severe psoriasis exhibited more pronounced metabolic alterations compared to those with non-severe disease. Although disease severity was defined only based on information within one year prior to blood sampling, since data on severity at the exact time of sample collection were unavailable, we found a higher GlycA elevation in severe psoriasis. This finding is consistent with previous research linking Psoriasis Area and Severity Index (PASI) score to GlycA.^34^

Furthermore, subgroup analysis indicated that the association between GlycA and psoriasis was largely driven by individuals with PsA, who exhibited higher GlycA levels. Interestingly, phenylalanine appeared to be selectively elevated in individuals with PsA, and not in those with PsC. In HUNT, this pattern was echoed in inflammatory arthritis, where elevated phenylalanine levels were also observed in RA and AS, but not in AD, UC, or CD. Although phenylalanine elevation was replicated only for RA in the UK Biobank, the associations may nonetheless be more specific to rheumatologic conditions, given that rheumatologic diagnoses were validated in HUNT but not in UK Biobank. Despite the limited number of studies reporting associations between phenylalanine and psoriasis or arthritis, our findings are supported by previous work identifying phenylalanine as a marker in osteoarthritis^38^ and showing elevated levels in diet-controlled RA cohorts.^39^

When we investigated other IMIDs, we found that systemic inflammation was present across all IMIDs, although its magnitude varied. In both cohorts, AD showed the least pronounced alterations, followed by PsC, suggesting that metabolomic disturbances in AD may be more localized to the skin and/or generally are less severe compared to psoriasis and other IMIDs. In HUNT, psoriasis showed a distinct metabolic pattern, primarily driven by lipoprotein subfractions. Although this pattern was not observed in the UK Biobank cohort, we emphasize the robustness of the psoriasis definition in HUNT. Most psoriasis cases in HUNT were based on self-reports; however, the psoriasis question has been formally validated by dermatologists, demonstrating a positive predictive value of 78%.^40^

To the best of our knowledge, this is the largest metabolomics study of psoriasis to date. Previous studies have been limited by small sample sizes, with most including fewer than 100 psoriasis cases, and few have validated their findings in independent cohorts.^6^ Adding further value, we also included other IMIDs, with case numbers among the largest, if not the largest, cohorts worldwide. In addition, in HUNT, all PsA, RA and AS cases were validated by rheumatologists^.15,16^

Current metabolomics studies in psoriasis have employed a wide variety of analytical platforms, making it challenging to compare results across studies. In our study, metabolites in both cohorts were measured using the same platform (Nightingale Health), thereby improving comparability. However, as our cohorts consisted predominantly of individuals of White ethnicity/European ancestry, the generalizability of our findings to other ethnic groups is limited and warrants further investigation in more diverse populations.

Despite efforts to enhance comparability, some differences between the two cohorts remain. First, the sample type differs: UK Biobank used EDTA plasma, whereas HUNT used serum, which may affect metabolite quantification,^41^ although the exact impact on our results is unclear. Second, UK Biobank had a much lower participation rate (5.5%), which may introduce selection bias toward healthier participants.^42^ In contrast, this was less of a concern in HUNT, which had a higher participation rate (54.1%).^43^ Third, differences in disease and covariate definitions also exist; for example, we used self-reported lipid-lowering medication use in UK Biobank versus prescription registry data in HUNT.

In conclusion, our study identified circulating metabolites associated with psoriasis, including cutaneous disease activity, severity, and the presence of PsA, highlighting systemic metabolic alterations linked to the disease. We also provide insights into the metabolic profile of psoriasis in the context of other IMIDs, revealing both shared and disease-specific metabolic features. Collectively, these findings enhance our understanding of psoriasis pathophysiology and provide a foundation for future metabolomics research.

## Acknowledgements

The Trøndelag Health Study (The HUNT Study) is a collaboration between HUNT Research Centre (Faculty of Medicine and Health Sciences, NTNU - Norwegian University of Science and Technology), Trøndelag County Council, Central Norway Regional Health Authority, and the Norwegian Institute of Public Health.

We thank UK Biobank and HUNT participants for donating their time, samples, and information to help others; clinicians and other employees at UK Biobank and Nord-Trøndelag Hospital Trust for their support and for contributing to data collection in this research project.

Data analyses were performed in digital laboratories at HUNT Cloud (NTNU - Norwegian University of Science and Technology, Trondheim, Norway) and UK Biobank Research Analysis Platform.

AI-assisted tools were used primarily for grammar checking, through Copilot, which is subscribed to by the institution. Its use is approved by NTNU - Norwegian University of Science and Technology, as stated in the institutional guideline: https://i.ntnu.no/wiki/-/wiki/English/Copilot.

## Funding sources

ML is funded by grants from the Liaison Committee for Education, Research and Innovation in Central Norway and the Joint Research Committee between St Olavs Hospital and the Faculty of Medicine and Health Sciences, NTNU - Norwegian University of Science and Technology.

## Conflict of interest

Nothing to declare.

## Data availability

The UK Biobank data is available to researchers for all types of health-related research that is in the public interest. Data are available for approved researchers through the UK Biobank data-access protocol. For accessing the resource, this project has been approved with application number 40135. For more information, see: https://www.ukbiobank.ac.uk/enable-your-research/apply-for-access.

The HUNT data is stored in the HUNT databank (https://www.ntnu.edu/hunt/databank), and is accessible for researchers approved by the Regional Committee for Medical and Health Research Ethics. Researchers from other countries are welcome to apply in cooperation with a Norwegian Principal Investigator. For more information, see: http://www.ntnu.edu/hunt/data.

## Ethics statements

The UK Biobank has been approved by the Northwest Multi-centre Research Ethics Committee (MREC) as a Research Tissue Bank (RTB) approval, granted in 2011 and renewed every 5 years. For more information, see: https://www.ukbiobank.ac.uk/learn-more-about-uk-biobank/about-us/ethics.

The use of HUNT data has been approved by the Regional Committee for Medical and Health Research Ethics (REK), Central Norway, with application number 27420.

## Notes

### Competing Interest Statement

The authors have declared no competing interest.

### Funding Statement

This research is funded by grants from the Liaison Committee for Education, Research and Innovation in Central Norway and the Joint Research Committee between St Olavs Hospital and the Faculty of Medicine and Health Sciences, NTNU - Norwegian University of Science and Technology.

### Author Declarations

The UK Biobank has been approved by the Northwest Multi-centre Research Ethics Committee (MREC) as a Research Tissue Bank (RTB) approval, granted in 2011 and renewed every 5 years. For more information, see: https://www.ukbiobank.ac.uk/learn-more-about-uk-biobank/about-us/ethics. The use of HUNT data has been approved by the Regional Committee for Medical and Health Research Ethics (REK), Central Norway, with application number 27420.

## References

1 Griffiths CEM, Armstrong AW, Gudjonsson JE et al. Psoriasis. Lancet 2021; 397: 1301–15.

2 Takeshita J, Grewal S, Langan SM et al. Psoriasis and comorbid diseases: Epidemiology. J. Am. Acad. Dermatol. 2017; 76: 377–90.

3 Reid C, Griffiths CEM. Psoriasis and Treatment: Past, Present and Future Aspects. Acta Derm. Venereol. 2020; 100: adv00032.

4 Mrowietz U, Kragballe K, Reich K et al. Definition of treatment goals for moderate to severe psoriasis: a European consensus. Arch. Dermatol. Res. 2011; 303: 1–10.

5 Reich K. The concept of psoriasis as a systemic inflammation: implications for disease management. J. Eur. Acad. Dermatol. Venereol. 2012; 26 Suppl 2: 3–11.

6 Koussiouris J, Looby N, Anderson M et al. Metabolomics Studies in Psoriatic Disease: A Review. Metabolites 2021; 11.

7 Guo L, Jin H. Research progress of metabolomics in psoriasis. Chin. Med. J. (Engl.) 2023; 136: 1805–16.

8 Ioannidis JPA, Bossuyt PMM. Waste, Leaks, and Failures in the Biomarker Pipeline. Clin. Chem. 2017; 63: 963–72.

9 Elliott P, Peakman TC, Biobank UK. The UK Biobank sample handling and storage protocol for the collection, processing and archiving of human blood and urine. Int. J. Epidemiol. 2008; 37: 234–44.

10 Sudlow C, Gallacher J, Allen N et al. UK biobank: an open access resource for identifying the causes of a wide range of complex diseases of middle and old age. PLoS Med. 2015; 12: e1001779.

11 Krokstad S, Langhammer A, Hveem K et al. Cohort Profile: the HUNT Study, Norway. Int. J. Epidemiol. 2013; 42: 968–77.

12 Asvold BO, Langhammer A, Rehn TA et al. Cohort Profile Update: The HUNT Study, Norway. Int. J. Epidemiol. 2023; 52: e80–e91.

13 Næss M, Kvaløy K, Sørgjerd EP et al. Data Resource Profile: The HUNT Biobank. Int. J. Epidemiol. 2024; 53.

14 Brumpton BM, Graham S, Surakka I et al. The HUNT study: A population-based cohort for genetic research. Cell Genom 2022; 2: 100193.

15 Hoff M, Gulati AM, Romundstad PR et al. Prevalence and incidence rates of psoriatic arthritis in central Norway: data from the Nord-Trondelag health study (HUNT). Ann. Rheum. Dis. 2015; 74: 60–4.

16 Videm V, Thomas R, Brown MA et al. Self-reported Diagnosis of Rheumatoid Arthritis or Ankylosing Spondylitis Has Low Accuracy: Data from the Nord-Trondelag Health Study. J. Rheumatol. 2017; 44: 1134–41.

17 Ramessur R, Dand N, Langan SM et al. Defining disease severity in atopic dermatitis and psoriasis for the application to biomarker research: an interdisciplinary perspective. Br. J. Dermatol. 2024; 191: 14–23.

18 Ramessur R, Gill D. The effect of statins on severity of psoriasis: A systematic review. Indian J. Dermatol. Venereol. Leprol. 2017; 83: 154–61.

19 Costanzo M, Caterino M, Sotgiu G et al. Sex differences in the human metabolome. Biol. Sex Differ. 2022; 13: 30.

20 Panyard DJ, Yu B, Snyder MP. The metabolomics of human aging: Advances, challenges, and opportunities. Sci Adv 2022; 8: eadd6155.

21 Budu-Aggrey A, Brumpton B, Tyrrell J et al. Evidence of a causal relationship between body mass index and psoriasis: A mendelian randomization study. PLoS Med. 2019; 16: e1002739.

22 Wei J, Zhu J, Xu H et al. Alcohol consumption and smoking in relation to psoriasis: a Mendelian randomization study. Br. J. Dermatol. 2022; 187: 684–91.

23 Zhao SS, Yiu ZZN, Barton A et al. Association of Lipid-Lowering Drugs With Risk of Psoriasis: A Mendelian Randomization Study. JAMA Dermatol 2023; 159: 275–80.

24 Soininen P, Kangas AJ, Wurtz P et al. Quantitative serum nuclear magnetic resonance metabolomics in cardiovascular epidemiology and genetics. Circ. Cardiovasc. Genet. 2015; 8: 192–206.

25 Ala-Korpela M, Zhao S, Jarvelin MR et al. Apt interpretation of comprehensive lipoprotein data in large-scale epidemiology: disclosure of fundamental structural and metabolic relationships. Int. J. Epidemiol. 2022; 51: 996–1011.

26 Ritchie SC, Surendran P, Karthikeyan S et al. Quality control and removal of technical variation of NMR metabolic biomarker data in ∼120,000 UK Biobank participants. Sci Data 2023; 10: 64.

27 Denos M, Sharma A, Åsvold BO et al. Associations between circulating metabolites and incidence of breast, prostate, lung and colorectal cancers in the HUNT Study and UK Biobank. medRxiv 2025.

28 Unione L, Arda A, Jimenez-Barbero J et al. NMR of glycoproteins: profiling, structure, conformation and interactions. Curr. Opin. Struct. Biol. 2021; 68: 9–17.

29 Crick DCP, Khandaker GM, Halligan SL et al. Comparison of the stability of glycoprotein acetyls and high sensitivity C-reactive protein as markers of chronic inflammation. Immunology 2024; 171: 497–512.

30 Connelly MA, Otvos JD, Shalaurova I et al. GlycA, a novel biomarker of systemic inflammation and cardiovascular disease risk. J. Transl. Med. 2017; 15: 219.

31 Solberg SM. Systemic inflammation in psoriasis: Circulating immune cells and cytokines. In: Department of Clinical Science, Vol. 2019: University of Bergen. 2019.

32 Visser MJE, Venter C, Roberts TJ et al. Psoriatic disease is associated with systemic inflammation, endothelial activation, and altered haemostatic function. Sci. Rep. 2021; 11: 13043.

33 Joshi AA, Lerman JB, Aberra TM et al. GlycA Is a Novel Biomarker of Inflammation and Subclinical Cardiovascular Disease in Psoriasis. Circ. Res. 2016; 119: 1242–53.

34 Svedbom A, Mallbris L, Gonzalez-Cantero A et al. Skin Inflammation, Systemic Inflammation, and Cardiovascular Disease in Psoriasis. JAMA Dermatol 2025; 161: 81–6.

35 Bartlett DB, Connelly MA, AbouAssi H et al. A novel inflammatory biomarker, GlycA, associates with disease activity in rheumatoid arthritis and cardio-metabolic risk in BMI-matched controls. Arthritis Res. Ther. 2016; 18: 86.

36 Chung CP, Ormseth MJ, Connelly MA et al. GlycA, a novel marker of inflammation, is elevated in systemic lupus erythematosus. Lupus 2016; 25: 296–300.

37 Francis L, Capon F, Smith CH et al. Inflammatory memory in psoriasis: From remission to recurrence. J. Allergy Clin. Immunol. 2024; 154: 42–50.

38 Zhai G, Sun X, Randell EW et al. Phenylalanine Is a Novel Marker for Radiographic Knee Osteoarthritis Progression: The MOST Study. J. Rheumatol. 2021; 48: 123–8.

39 Lindqvist HM, Gjertsson I, Hulander E et al. Exploring the differences in serum metabolite profiles after intake of red meat in women with rheumatoid arthritis and a matched control group. Eur. J. Nutr. 2024; 63: 221–30.

40 Modalsli EH, Snekvik I, Asvold BO et al. Validity of Self-Reported Psoriasis in a General Population: The HUNT Study, Norway. J. Invest. Dermatol. 2016; 136: 323–5.

41 Vignoli A, Tenori L, Morsiani C et al. Serum or Plasma (and Which Plasma), That Is the Question. J. Proteome Res. 2022; 21: 1061–72.

42 Fry A, Littlejohns TJ, Sudlow C et al. Comparison of Sociodemographic and Health-Related Characteristics of UK Biobank Participants With Those of the General Population. Am. J. Epidemiol. 2017; 186: 1026–34.

43 Langhammer A, Krokstad S, Romundstad P et al. The HUNT study: participation is associated with survival and depends on socioeconomic status, diseases and symptoms. BMC Med. Res. Methodol. 2012; 12: 143.

