## Supplementary Data for "Circulating metabolites associated with psoriasis in the UK Biobank and the HUNT Study: A cross-sectional study of 270,848 participants"

### Supplementary Figures

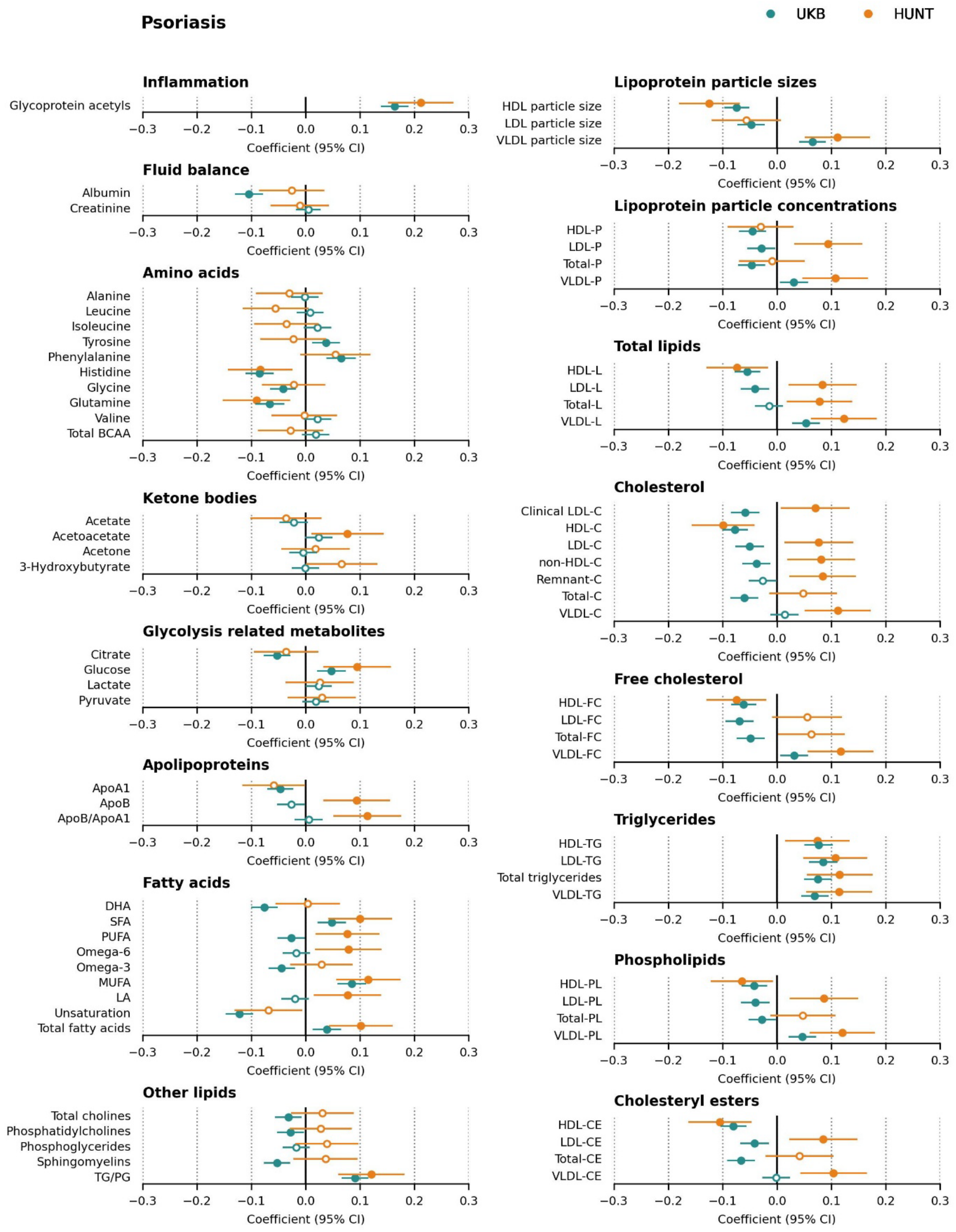

**Supplementary Figure S1:** Associations between small-molecule metabolites, fatty acids, and lipoprotein classes with psoriasis. The linear model was adjusted for age and sex (model 1). Adjusted p-values < 0.05 are indicated by solid fill. The forest plot shows only the absolute measures; the full results, including ratio measures, are provided in the **Supplementary Table S1.** The list of metabolite abbreviations is provided in **Supplementary Table S8.** UKB = UK Biobank, CI = Confidence Interval.

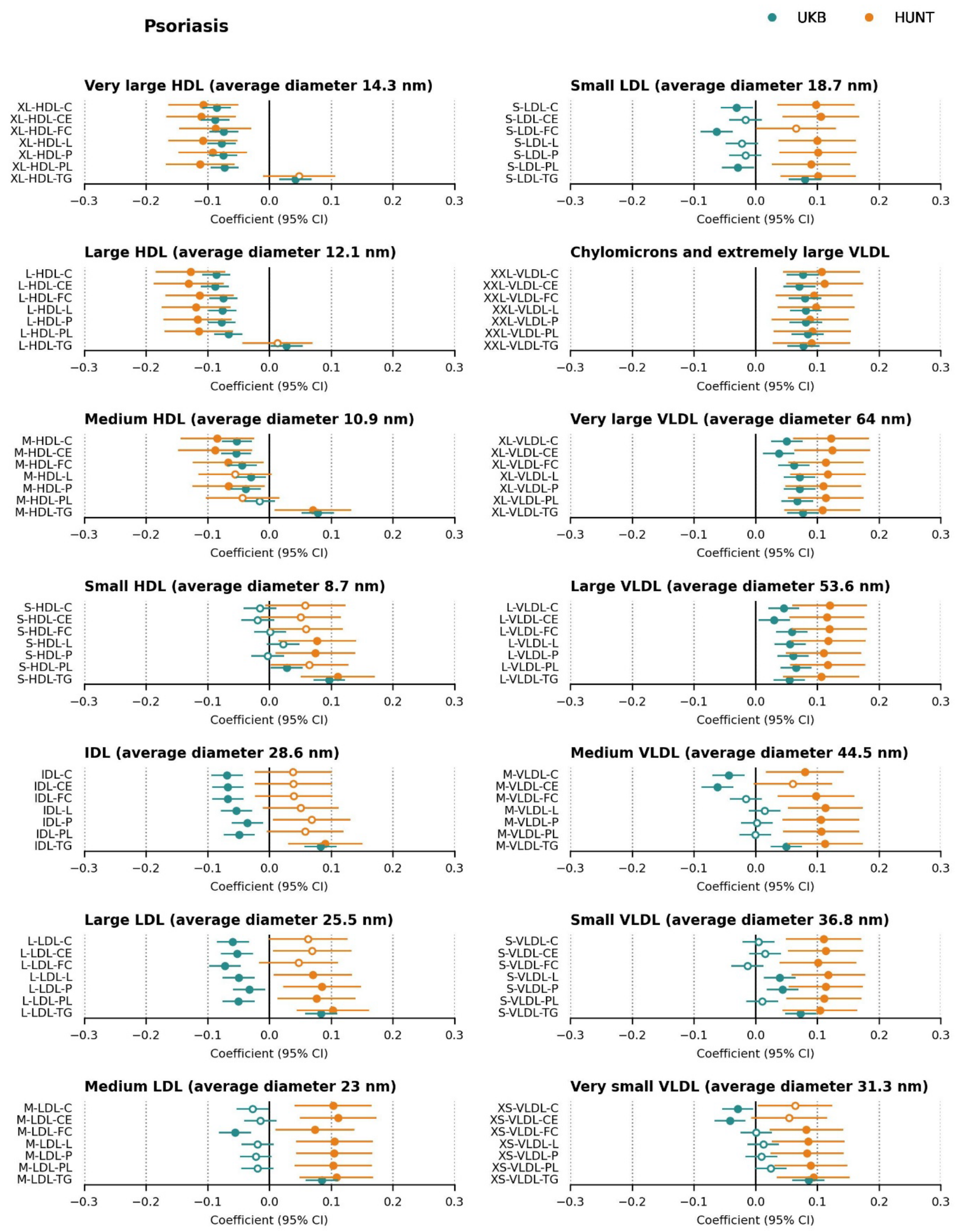

**Supplementary Figure S1 (continued):** Associations between lipoprotein sub-classes and sub-fractions with psoriasis. The linear model was adjusted for age and sex (model 1). Adjusted p-values < 0.05 are indicated by solid fill. The forest plot shows only the absolute measures; the full results, including ratio measures, are provided in the **Supplementary Table S1.** The list of metabolite abbreviations is provided in **Supplementary Table S8.** UKB = UK Biobank, CI = Confidence Interval.

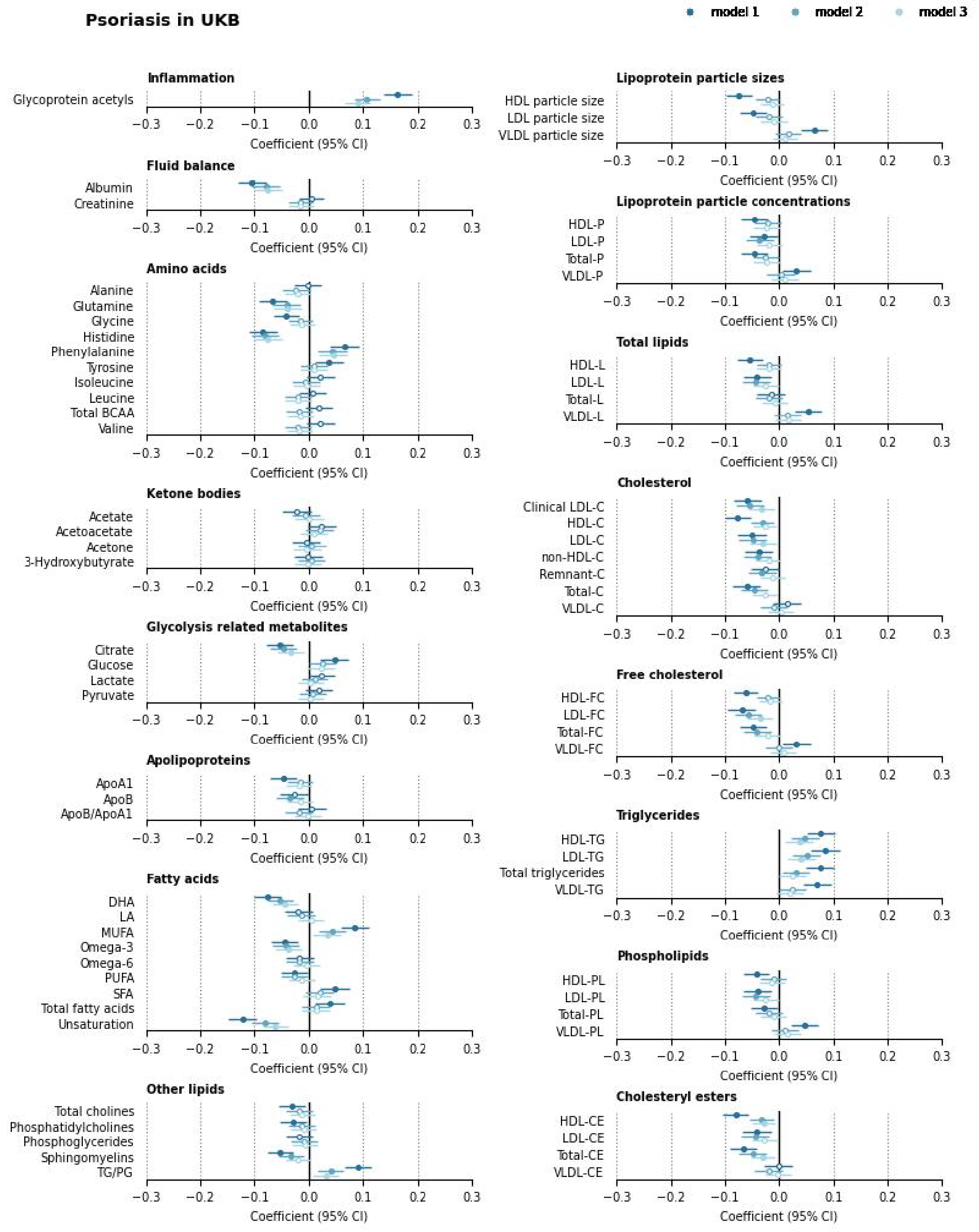

**Supplementary Figure S2:** Associations between small molecule metabolites, fatty acids, and lipoprotein classes with psoriasis in UK Biobank, using models 1, 2, and 3. Model 1 is adjusted for age and sex; Model 2 for age, sex, and BMI; and Model 3 for age, sex, BMI, smoking status, and use of lipid-lowering medication. Adjusted p-values < 0.05 are indicated by solid fill. The forest plot shows only the absolute measures; the full results, including ratio measures, are provided in the **Supplementary Table S1**. The list of metabolite abbreviations is provided in **Supplementary Table S8.** CI = Confidence Interval.

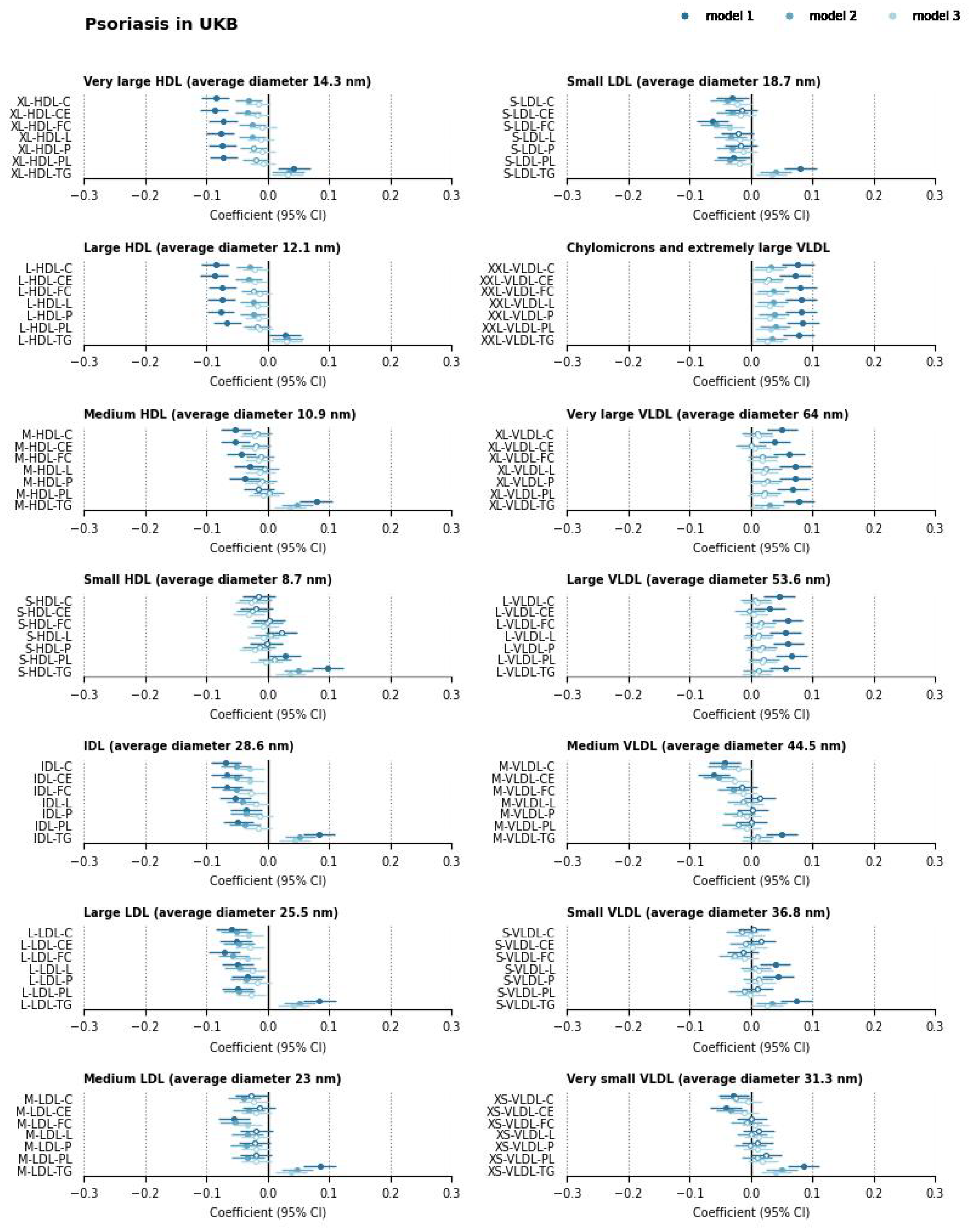

**Supplementary Figure S2 (continued):** Associations between lipoprotein sub-classes and sub-fractions with psoriasis in UK Biobank, using models 1, 2, and 3. Model 1 is adjusted for age and sex; Model 2 for age, sex, and BMI; Model 3 for age, sex, BMI, smoking status, and use of lipid-lowering medication. Adjusted p-values < 0.05 are indicated by solid fill. The forest plot shows only the absolute measures; the full results, including ratio measures, are provided in the **Supplementary Table S1**. The list of metabolite abbreviations is provided in **Supplementary Table S8.** CI = Confidence Interval.

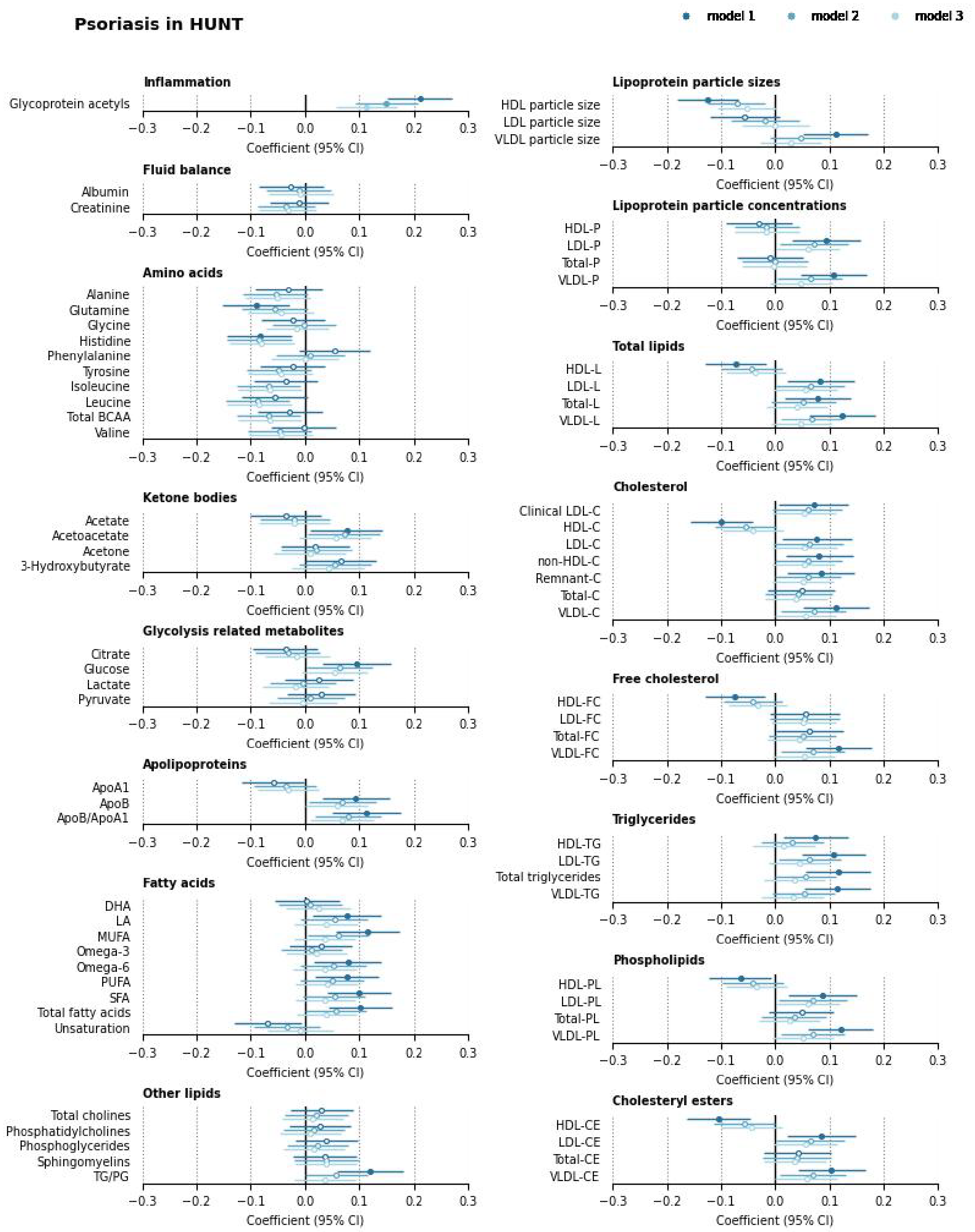

**Supplementary Figure S3:** Associations between small molecule metabolites, fatty acids, and lipoprotein classes with psoriasis in HUNT, using models 1, 2, and 3. Model 1 is adjusted for age and sex; Model 2 for age, sex, and BMI; Model 3 for age, sex, BMI, smoking status, and use of lipid-lowering medication. Adjusted p-values < 0.05 are indicated by solid fill. The forest plot shows only the absolute measures; the full results, including ratio measures, are provided in the **Supplementary Table S1**. The list of metabolite abbreviations is provided in **Supplementary Table S8.** CI = Confidence Interval.

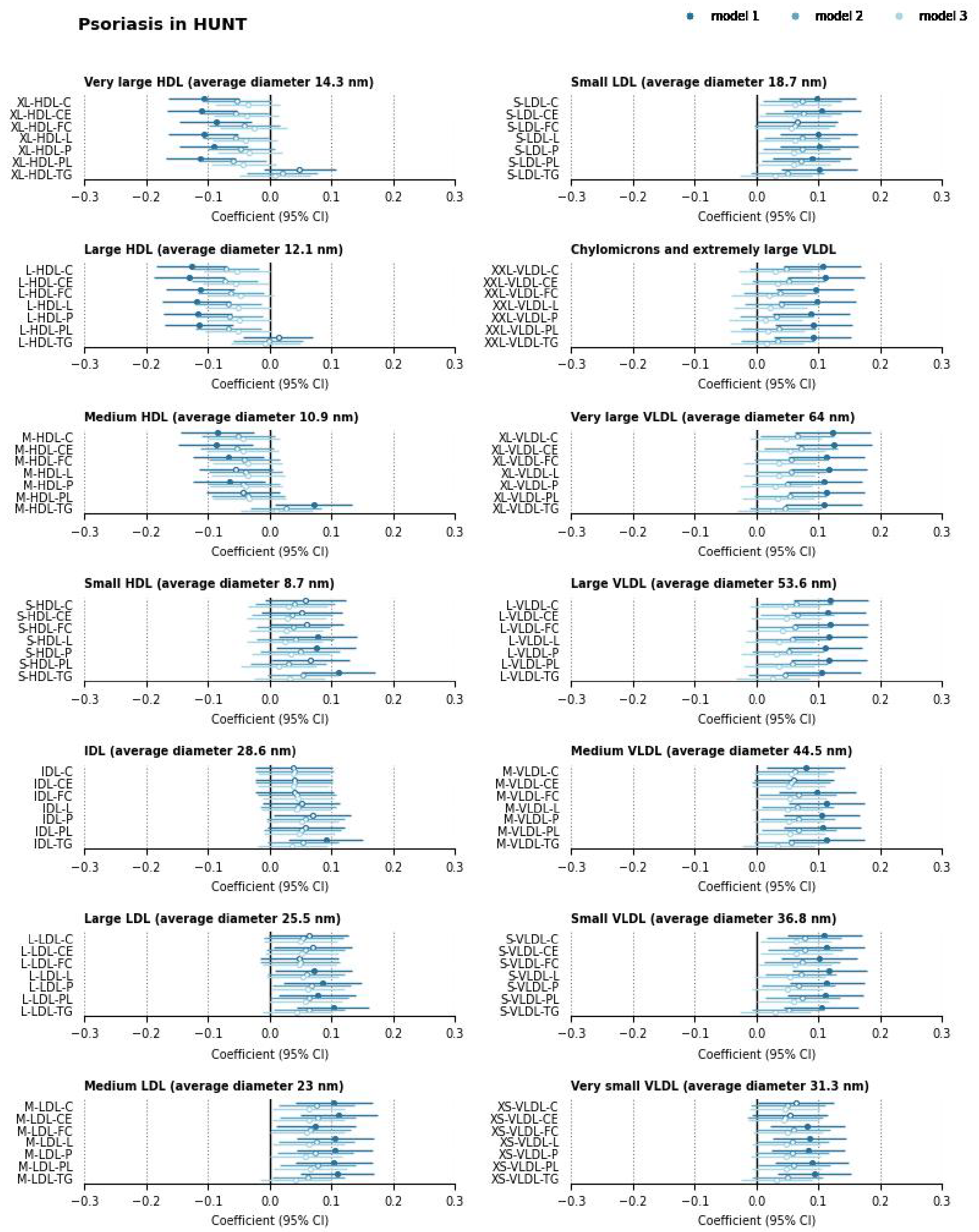

**Supplementary Figure S3 (continued):** Associations between lipoprotein sub-classes and sub-fractions with psoriasis in HUNT, using models 1, 2, and 3. Model 1 is adjusted for age and sex; Model 2 for age, sex, and BMI; Model 3 for age, sex, BMI, smoking status, and use of lipid-lowering medication. Adjusted p-values < 0.05 are indicated by solid fill. The forest plot shows only the absolute measures; the full results, including ratio measures, are provided in the **Supplementary Table S1**. The list of metabolite abbreviations is provided in **Supplementary Table S8.** CI = Confidence Interval.

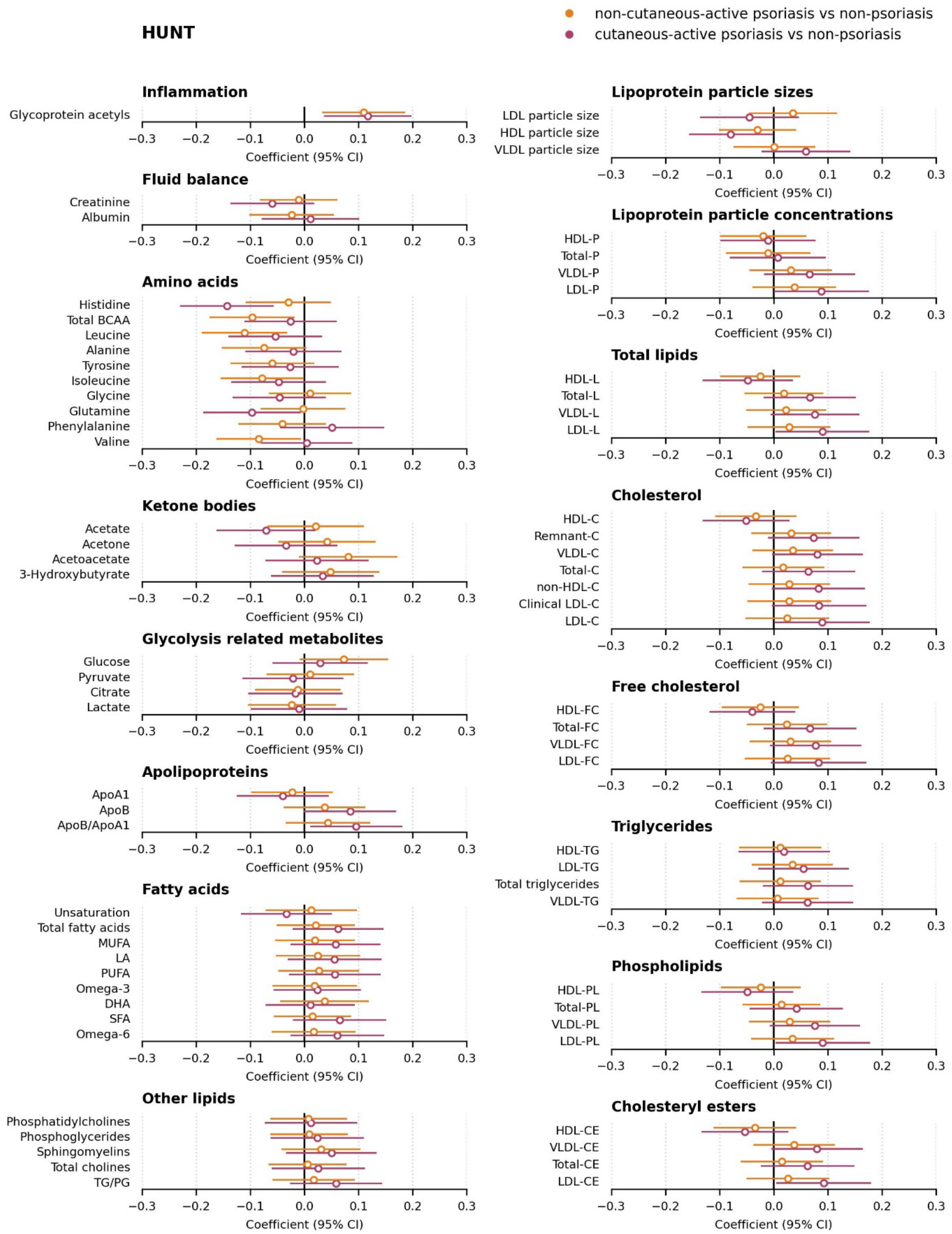

**Supplementary Figure S4:** Associations between small molecule metabolites, fatty acids, and lipoprotein classes with non-cutaneous-active and cutaneous-active psoriasis in HUNT, in the fully adjusted model (Model 3). Adjusted p-values < 0.05 are indicated by solid fill. The forest plot shows only the absolute measures; the full results, including ratio measures, are provided in the **Supplementary Table S3**. The list of metabolite abbreviations is provided in **Supplementary Table S8.** CI = Confidence Interval.

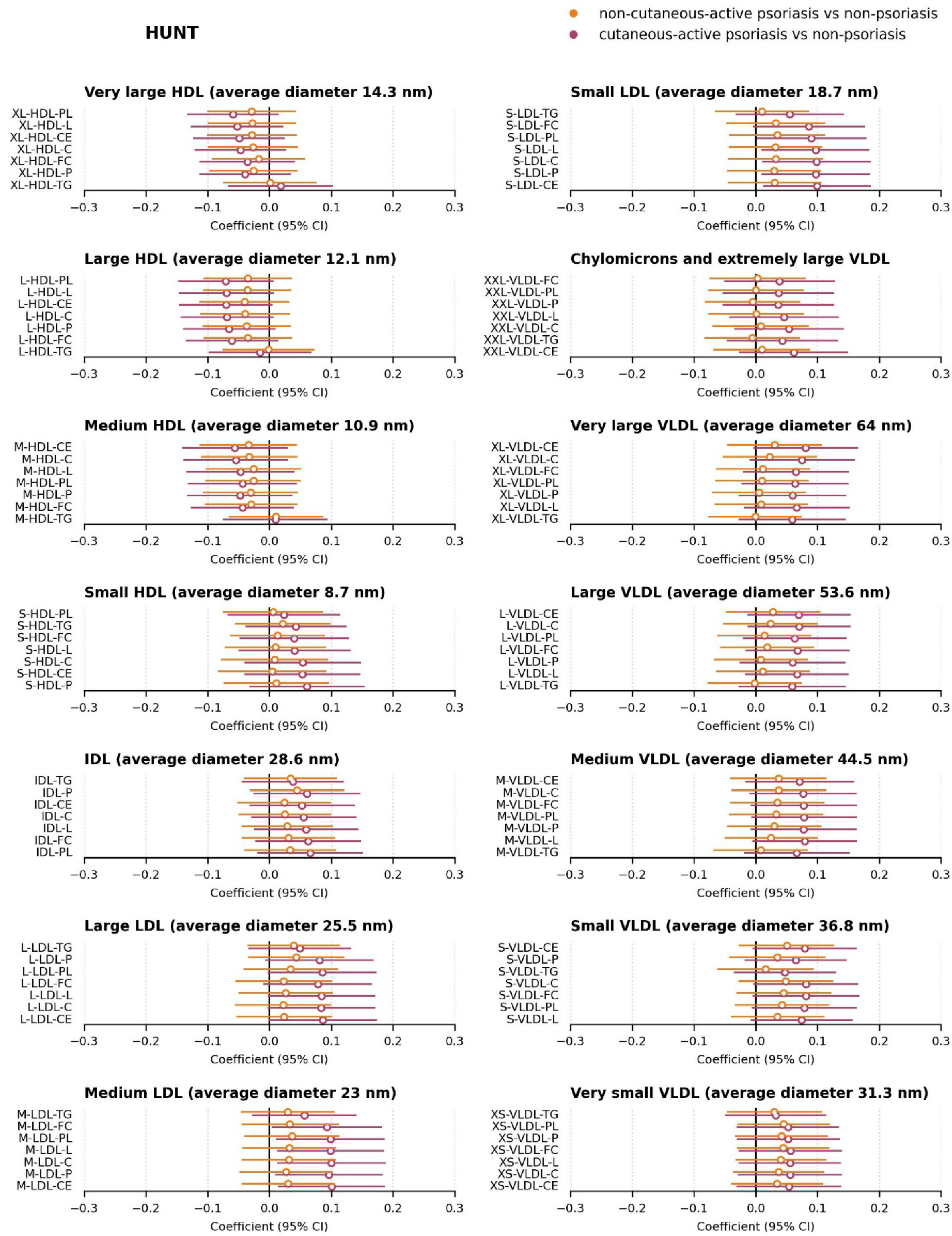

**Supplementary Figure S4 (continued):** Associations between lipoprotein sub-classes and sub-fractions with non-cutaneous-active psoriasis and cutaneous-active psoriasis in HUNT, in the fully adjusted model (Model 3). Adjusted p-values < 0.05 are indicated by solid fill. The forest plot shows only the absolute measures; the full results, including ratio measures, are provided in the **Supplementary Table S3.** The list of metabolite abbreviations is provided in **Supplementary Table S8.** CI = Confidence Interval.

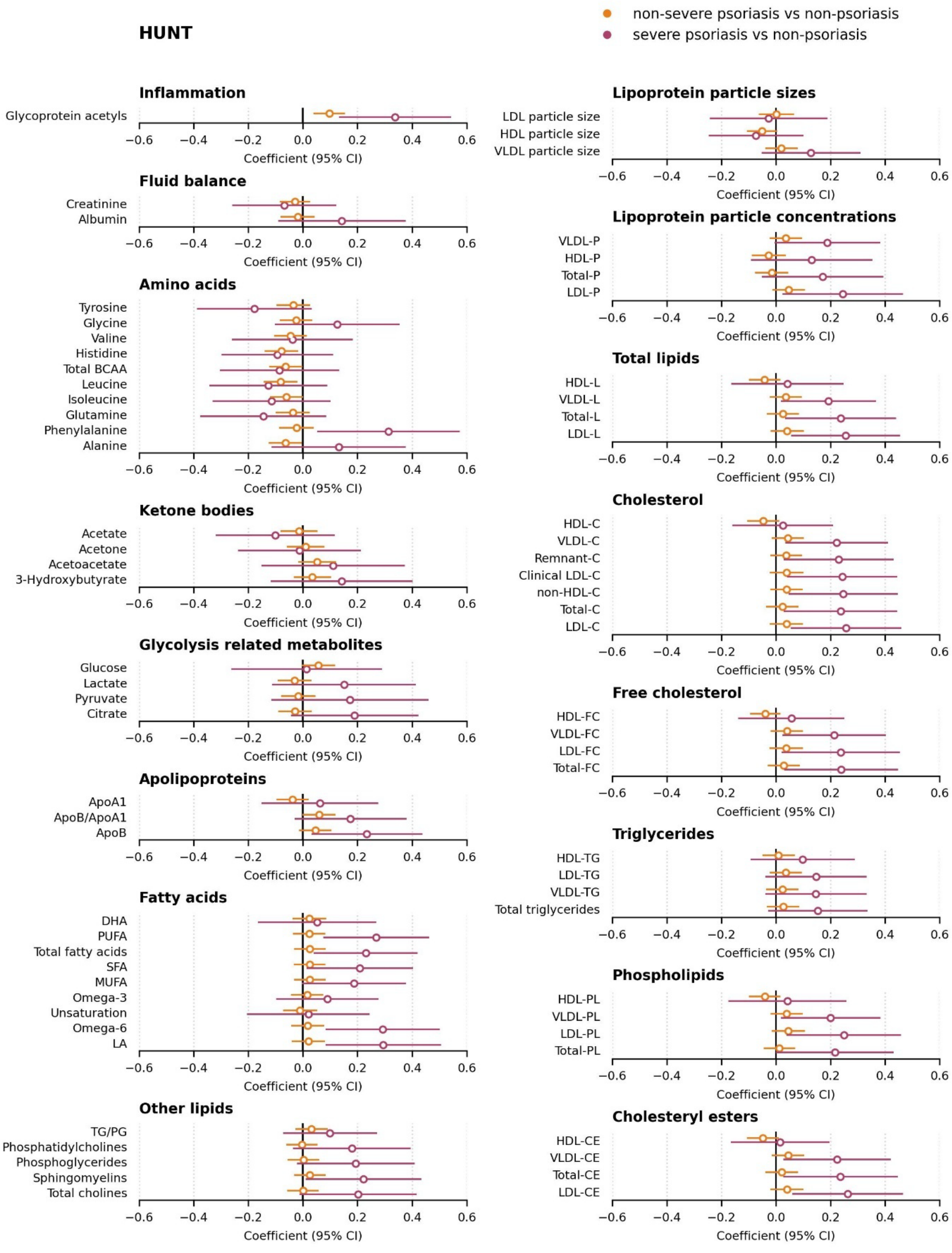

**Supplementary Figure S5:** Associations between small molecule metabolites, fatty acids, and lipoprotein classes with non-severe and severe active psoriasis in HUNT, in the fully adjusted model (Model 3). Adjusted p-values < 0.05 are indicated by solid fill. The forest plot shows only the absolute measures; the full results, including ratio measures, are provided in the **Supplementary Table S4**. The list of metabolite abbreviations is provided in **Supplementary Table S8.** CI = Confidence Interval.

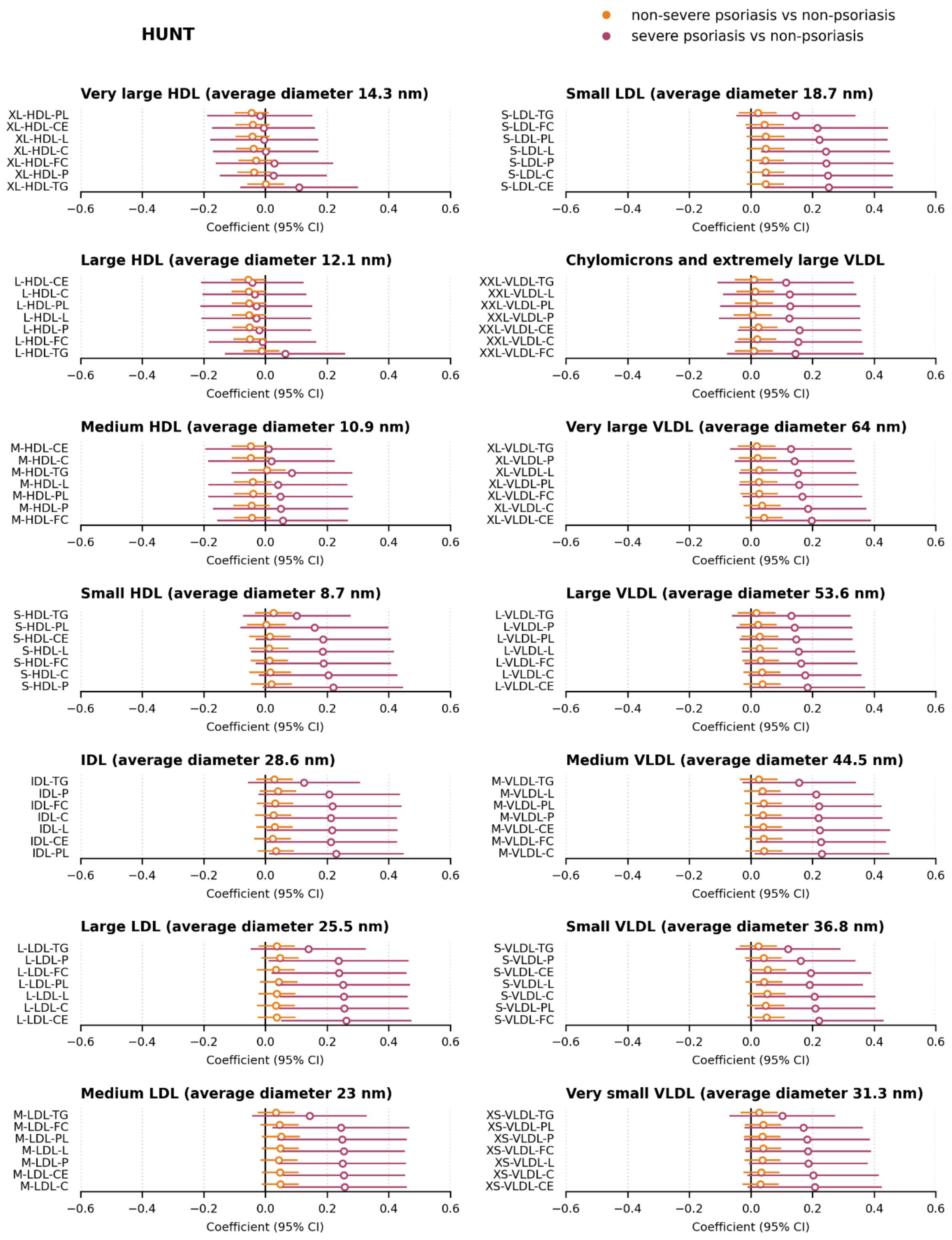

**Supplementary Figure S5 (continued):** Associations between lipoprotein sub-classes and sub-fractions with non-severe and severe active psoriasis in HUNT, in the fully adjusted model (Model 3). Adjusted p-values < 0.05 are indicated by solid fill. The forest plot shows only the absolute measures; the full results, including ratio measures, are provided in the **Supplementary Table S4**. The list of metabolite abbreviations is provided in **Supplementary Table S8.** CI = Confidence Interval.

### Supplementary Methods

**Supplementary Method S1.** Psoriasis case definitions in UK Biobank and HUNT.

| | UK Biobank | HUNT |
|---|---|---|
| Psoriasis | Individuals diagnosed with any type of psoriasis (ICD-10 code L40), defined by the date of psoriasis first occurrence (data field 131742) relative to the blood sampling (date of attendance at the assessment center, data field 53).<br><br>Only individuals diagnosed prior to blood sampling were included. | Individuals self-reporting psoriasis in the HUNT3 baseline questionnaire by answering *"Yes"* to the question: *"Have you had, or do you have psoriasis?"* (variable: PsorEv@NT3BLQ1)<br><br>OR<br><br>Individuals with any of the following ICD codes on their medical records (Helse Nord-Trøndelag (HNT) database):<br>ICD-9 codes: 696.1, 696.10, 696.12<br>ICD-10 codes: L40.0, L40.1, L40.2, L40.3, L40.4, L40.8, L40.9<br><br>OR<br><br>Individuals with any of the following codes in the Norwegian Control and Payment of Health Reimbursement (KUHR) database:<br>ICD-10 codes: L40, L40.0, L40.1, L40.2, L40.3, L40.4, L40.8, L40.9<br>ICPC-2 codes: S91, S910002, S910005<br><br>OR<br><br>Individuals with any prescriptions for psoriasis (ICD-10: L40; ICPC-2: S91) in the Norwegian Prescribed Drug Registry.<br><br>Only individuals with a first diagnosis date recorded in HNT or KUHR, or a first prescription date in the Norwegian Prescribed Drug Registry prior to blood sampling (variable: PartDat@NT3BLQ1) were included. |
| Cutaneous-active psoriasis | Not available. | Defined based on the HUNT3-Q3 psoriasis questionnaire, using the following criteria:<br><br>Individuals answering *"Yes"* to the question, *"Do you have a psoriasis rash at the moment?"* (variable: PsorCu@NT3Der1Q2)<br><br>OR<br><br>Individuals who reported a psoriasis outbreak in the past 14 days (variable: PsorLocL2W@NT3Der1Q2). |

| | | |
|---|---|---|
| Severe psoriasis (within one year prior to blood sampling) | Not available due to insufficient information to infer psoriasis severity within one year to blood sampling. | Severity criteria were adapted from previous literature (Ramessur et al., 2024). Cases were defined as severe psoriasis occurring within one year prior to blood sampling, identified using the following criteria:<br><br>Individuals answering "*Yes*" to the question "*Have you ever taken tablets or injections for psoriasis*" in the HUNT3-Q3 psoriasis questionnaire (Variable: TreTabInjEv@NT3Der1Q2). If "*Yes*": "*Have you, in the last 12 months or the last 14 days, taken any of the following medicines (Methotrexate / Neotigason / Sandimmune / PUVA treatment / Embrel / Remicade / Raptiva / Humira / Other) for psoriasis?*"<br><br>OR<br><br>Individuals with hospital admission (inpatient clinics) due to psoriasis (main diagnosis):<br>ICD-9: 696.1, 696.10, 696.12<br>ICD-10: L40, L40.0, L40.1, L40.2, L40.3, L40.4, L40.8, L40.9<br><br>OR<br><br>Individuals with phototherapy treatment defined by any of the NCMP-NCSP-NCRP codes: QXGX00, QXGX10, QXGX25, QXGX30 given for ICD-10: L40, L40.0, L40.1, L40.2, L40.3, L40.4, L40.8, L40.9 in the Norwegian Control and Payment of Health Reimbursement (KUHR) database<br><br>OR<br><br>Individuals with systemic medication in the Norwegian Prescribed Drug Registry for ICD-10 L40 or ICPC-2 S91 on ATC codes indicating systemic therapies: L04AX03, L01BA01, D05BB02, L04AX07, L04AD01, D05BX51, D05BA03, L04AB04, L04AB05, L04AB01, L04AB02, L04AA21, L04AC12, L04AC13, L04AC10, L04AC21, L04AC17, L04AC16, L04AC18, L04AC05, L04AA56, or D05BA02. |
| Psoriatic arthritis | Individuals with psoriasis were diagnosed with psoriatic arthritis (ICD-10 codes L40.5, M07.0, M07.1, M07.2, or M07.3), defined by the summary diagnosis (data fields 41270 and 41280) relative to the blood sampling date (date of attendance at the assessment center, data field 53).<br><br>Only individuals diagnosed prior to sampling were included. | Individuals self-reporting psoriatic arthritis in HUNT by answering *"Yes"* to the questionnaire question, *"Do you have psoriatic arthritis?"* (variable: PsorArth@NT3Der1Q2), which was subsequently verified by rheumatologists (Hoff et al., 2015). |

**Supplementary Method S2.** Other IMIDs definitions

Currently, there is no formal definition or comprehensive list of immune-mediated inflammatory diseases (IMIDs) in the literature. In this study, we selected several IMIDs primarily based on their shared immunopathogenic pathways (Schett et al., 2021) and the number of available cases in our datasets. The definitions used for each IMID are provided in table below.

| | UK Biobank | HUNT |
|---|---|---|
| Atopic dermatitis | Individuals diagnosed with ICD-10 code L20, defined by the date of first occurrence (data field 131720) relative to the blood sampling date (date of attendance at the assessment center, data field 53).<br><br>Only individuals diagnosed prior to sampling were included. | Individuals self-reporting eczema in HUNT by answering *"Yes"* to the questionnaire question, *"Have you had, or do you have eczema (on hands)?"* (variable: EczEv@NT3BLQ1)<br><br>OR<br><br>Individuals with any of the following ICD codes on the medical records (HNT database):<br>ICD-9 codes: 691.8*<br>ICD-10 codes: L20*<br><br>OR<br><br>Individuals with any of the following codes in the billing records (KUHR database):<br>ICD-10 codes: L20*<br>ICPC-2 codes: S87*<br><br>OR<br><br>Individuals with any prescriptions for psoriasis (ICD-10: L20* and ICPC-2: S87*) in the Norwegian Prescribed Drug Registry.<br><br>Only individuals with a first diagnosis date from HNT or KUHR, or a first prescription date from Norwegian Prescribed Drug Registry prior to the sampling time (variable: PartDat@NT3BLQ1) were included. |
| Rheumatoid arthritis | Individuals diagnosed with ICD-10 codes M05 and M06, defined by the date of first occurrence (data fields 131848 and 131850) relative to the blood sampling date (date of attendance at the assessment center, data field 53).<br><br>Only individuals diagnosed prior to sampling were included. | Individuals self-reporting rheumatoid arthritis in HUNT by answering *"Yes"* to the questionnaire question, *"Have you had, or do you have arthritis (rheumatoid arthritis)?"* (variable: RhArthEv@NT3BLQ1), which were subsequently verified by rheumatologists (Videm et al., 2017). |
| Ankylosing spondylitis | Individuals diagnosed with ICD-10 code M45, defined by the date of first occurrence (data field 131912) relative to the blood sampling date (date attendance at the assessment | Individuals self-reporting spondyloarthritis in HUNT by answering *"Yes"* to the questionnaire question, *"Have you had, or do you have Bechterew's disease?"* (variable: SponArthEv@NT3BLQ1), which were subsequently verified by rheumatologists (Videm et al., 2017). |

| | | |
|---|---|---|
| | center, data field 53).<br><br>Only individuals diagnosed prior to sampling were included. | |
| Systemic lupus erythematosus | Individuals diagnosed with ICD-10 code M32, defined by the date of first occurrence (data field 131894) relative to the blood sampling date (date of attendance at the assessment center, data field 53).<br><br>Only individuals diagnosed prior to sampling were included. | Individuals with any of the following ICD codes on the medical records (HNT database):<br>ICD-9 code: 710.0<br>ICD-10 codes: M32*<br><br>OR<br><br>Individuals with any of the following codes on the billing records (KUHR database):<br>ICD-10 codes: M32*<br><br>OR<br><br>Individuals with any prescriptions for SLE (ICD-10: M32*) in the Norwegian Prescribed Drug Registry. |
| Ulcerative colitis | Individuals diagnosed with ICD-10 code K51, defined by the date of first occurrence (data field 131628) relative to the blood sampling date (date of attendance at the assessment center, data field 53).<br><br>Only individuals diagnosed prior to sampling were included. | Individuals with any of the following ICD codes on the medical records (HNT database):<br>ICD-9 codes: 556*<br>ICD-10 codes: K51*<br><br>OR<br><br>Individuals with any of the following codes on the billing records (KUHR database):<br>ICD-10 codes: K51*<br><br>OR<br><br>Individuals with any prescriptions for UC (ICD-10: K51*) in the Norwegian Prescribed Drug Registry. |
| Crohn's disease | Individuals diagnosed with ICD-10 code K50, defined by the date of first occurrence (data field 131626) relative to the blood sampling date (date of attendance at the assessment center, data field 53).<br><br>Only individuals diagnosed prior to sampling were included. | Individuals with any of the following ICD codes on the medical records (HNT database):<br>ICD-9 codes: 555*<br>ICD-10 codes: K50*<br><br>OR<br><br>Individuals with any of the following codes on the billing records (KUHR database):<br>ICD-10 codes: K50*<br><br>OR<br><br>Individuals with any prescriptions for CD (ICD-10: K50*) in the Norwegian Prescribed Drug Registry. |

**Supplementary Method S3.** UK Biobank data fields and HUNT variables used as covariates in the study.

| | UK Biobank data field | HUNT variable |
|---|---|---|
| Age | 21003 | PartAg@NT3BLQ1 |
| Sex | 31 | Sex |
| Ethnicity | 21000 | Defined from genetic data as described in previous literature (Brumpton et al., 2022) |
| BMI | 21001 | BMI@NT3BLM |
| Fasting time | 74 | TmLMe@NT3BLM |
| Smoking status | 20116 | SmoStat@NT3BLQ1 |
| Use of lipid-lowering medication | 6177 (male) and 6153 (female) | Defined as any participants with ATC code C10 within one year prior to the participation date, based on data from the Norwegian Prescribed Drug Registry. |

**Supplementary Method S4.** Metabolomics data imputation

Metabolites with missing values presumed to be missing not at random (MNAR)—likely due to spectrometer detection limits—were imputed using the Quantile Regression Imputation of Left-Censored data (QRILC) method, which has been shown to perform best for MNAR metabolomics data (Wei et al., 2018). In the UK Biobank, MNAR was inferred from the quality control (QC) flag "*limit of quantification*". In HUNT, all zero and missing values were assumed to fall below the detection threshold and were therefore treated as MNAR, due to the absence of QC flags as in the UK Biobank. For other types of missing values in the UK Biobank, we imputed using k-Nearest Neighbor (kNN) method, which offers strong performance while being less computationally intensive than more advanced approaches (Wei et al., 2018).

**References**

Brumpton, B. M., Graham, S., Surakka, I., Skogholt, A. H., Loset, M., Fritsche, L. G., Wolford, B., Zhou, W., Nielsen, J. B., Holmen, O. L., Gabrielsen, M. E., Thomas, L., Bhatta, L., Rasheed, H., Zhang, H., Kang, H. M., Hornsby, W., Moksnes, M. R., Coward, E.,…Willer, C. J. (2022). The HUNT study: A population-based cohort for genetic research. *Cell Genom*, *2*(10), 100193. https://doi.org/10.1016/j.xgen.2022.100193

Hoff, M., Gulati, A. M., Romundstad, P. R., Kavanaugh, A., & Haugeberg, G. (2015). Prevalence and incidence rates of psoriatic arthritis in central Norway: data from the Nord-Trondelag health study (HUNT). *Annals of the Rheumatic Diseases*, *74*(1), 60–64. https://doi.org/10.1136/annrheumdis-2013-203862

Ramessur, R., Dand, N., Langan, S. M., Saklatvala, J., Fritzsche, M. C., Holland, S., Arents, B. W. M., McAteer, H., Proctor, A., McMahon, D., Greenwood, M., Buyx, A. M., Messer, T., Weiler, N., Hicks, A., Hecht, P., Weidinger, S., Ndlovu, M. N., Chengliang, D.,…Smith, C. H. (2024). Defining disease severity in atopic dermatitis and psoriasis for the application to biomarker research: an interdisciplinary perspective. *British Journal of Dermatology*, *191*(1), 14–23. https://doi.org/10.1093/bjd/ljae080

Saklatvala, J. R., Lessard, S., Teder-Laving, M., Thomas, L. F., Ramessur, R., Åsvold, B. O., Barton, A., Baudry, D., Bowes, J., Brumpton, B., Chandran, V., Chatelain, C., de Rinaldis, E., Elder, J. T., Ellinghaus, D., Foerster, J., Franke, A., Gladman, D. D., Gulliver, W.,…Simpson, M. A. (2025). Genetic liability to psoriasis predicts severe disease outcomes. *medRxiv*. https://doi.org/10.1101/2025.03.04.25323079

Schett, G., McInnes, I. B., & Neurath, M. F. (2021). Reframing Immune-Mediated Inflammatory Diseases through Signature Cytokine Hubs. *New England Journal of Medicine*, *385*(7), 628–639. https://doi.org/10.1056/NEJMra1909094

Videm, V., Thomas, R., Brown, M. A., & Hoff, M. (2017). Self-reported Diagnosis of Rheumatoid Arthritis or Ankylosing Spondylitis Has Low Accuracy: Data from the Nord-Trondelag Health Study. *Journal of Rheumatology*, *44*(8), 1134–1141. https://doi.org/10.3899/jrheum.161396

Wei, R., Wang, J., Su, M., Jia, E., Chen, S., Chen, T., & Ni, Y. (2018). Missing Value Imputation Approach for Mass Spectrometry-based Metabolomics Data. *Scientific Reports*, *8*(1), 663. https://doi.org/10.1038/s41598-017-19120-0
